# Human IL-34 Deficiency Primes Microglia Toward Alzheimer’s Disease-Associated States

**DOI:** 10.64898/2026.02.21.26346696

**Authors:** Francisco Hernandez-Rasco, Rocío Ruiz, Itziar de Rojas, Raquel Puerta, Clara Garcia-Mayor, Ana María Espinosa-Oliva, Juan García-Revilla, Paula Bayon, Alberto Rivera-Ramos, Farah Real Oualit, Sebastián Jiménez, Maria Eugenia Saez, Rocío M. de Pablos, Feiyang Zhao, Claudia Olive, Pilar Sanz, Xavier Montalbán, Sergi Valero, Amanda Cano, María Victoria Fernández, Anne Marie Wells, Jose Enrique Cavazos, Sudha Seshadri, Merce Boada, Santos Mañes, Michael T. Heneka, Francisco Javier Vitorica, Alfredo Ramirez, José Luis Venero, Agustín Ruiz

**Affiliations:** Instituto de Biomedicina de Sevilla, IBIS/Hospital Universitario Virgen del Rocío/CSIC/Universidad de Sevilla, Sevilla, Spain; Departamento de Bioquímica y Biología Molecular, Facultad de Farmacia, Universidad de Sevilla, Sevilla, Spain; Research Center and Memory Clinic. Ace Alzheimer Center Barcelona- Universitat Internacional de Catalunya, Barcelona, Spain; Luxembourg Centre for Systems Biomedicine (LCSB), University of Luxembourg, Esch-sur-Alzette, Luxembourg; Doctorate in Biotechnology, Facultat de Farmàcia i Ciències de l’Alimentació, Universitat de Barcelona, Avda. Diagonal 643, 08028, Barcelona, Spain; Centro de Investigación Biomédica en Red sobre Enfermedades Neurodegenerativas (CIBERNED), Madrid, Spain; CAEBi, Centro Andaluz de Estudios Bioinformáticos, Sevilla, Spain; Glenn Biggs Institute for Alzheimer’s & Neurodegenerative Diseases, University of Texas at San Antonio, San Antonio, TX, USA; Department of Neurology, Multiple Sclerosis Centre of Catalonia (Cemcat), Hospital Universitari Vall d’Hebron, Universitat Autònoma de Barcelona, Barcelona, Spain; South Texas Alzheimer’s Disease Research Center, San Antonio, TX, USA; Department of Neurology. Long School of Medicine, University of Texas at San Antonio, San Antonio, Texas, USA; Department of Immunology and Oncology, Centro Nacional de Biotecnología/Consejo Superior de Investigaciones Científicas, Madrid, Spain; Division of Neurogenetics and Molecular Psychiatry, Department of Psychiatry and Psychotherapy, Faculty of Medicine and University Hospital Cologne, University of Cologne, Cologne, Germany; Department of Old Age Psychiatry and Cognitive Disorders, University Hospital Bonn, Bonn, Germany; Cologne Excellence Cluster on Cellular Stress Responses in Aging-Associated Diseases (CECAD), University of Cologne, Cologne, Germany; Department of Psychiatry. Long School of Medicine, University of Texas at San Antonio, San Antonio, Texas, USA; Department of Microbiology, Immunology and Molecular Genetics. Long School of Medicine, University of Texas at San Antonio, San Antonio, Texas, USA

## Abstract

**Background:** Genome-wide association studies (GWAS), with independent replication in large European consortia, have identified a common nonsense variant in IL-34 (Y213X) as a genetic risk factor for late-onset Alzheimer’s disease (AD). However, the biological consequences of this IL-34 mutation in humans, its prevalence in the population, and the mechanisms by which IL-34-Y213X alters microglial homeostasis, cerebrospinal fluid (CSF) proteomic networks, and amyloid pathology remain poorly understood.

**Methods:** We combined human genetics, cerebrospinal fluid (CSF) and serum proteomics, transcriptomics, large-scale phenome-wide association analyses, and preclinical experimental models to define the impact of human IL-34 deficiency. IL-34 concentrations were first quantified in CSF and serum from deeply phenotyped AD cohorts stratified by the common IL-34-Y213X nonsense variant. IL-34 levels and IL-34-Y213X status were then integrated with unbiased CSF proteomic networks and AD biomarkers. Transcriptomic profiling of purified microglia from IL-34 knockout mice was performed to assess disease-associated microglial programs. Using APP/PS1 mice lacking IL-34, we examined the effects of IL-34 deficiency on microglial survival, tiling, and plaque encapsulation. Finally, we performed postmortem analyses of temporal cortex from AD patients carrying IL-34-Y213X to assess microglial density, spatial organization, and plaque-associated responses.

**Findings:** IL-34-Y213X was a strong, dose-dependent loss-of-function (LOF) allele that reduced IL-34 levels by up to 2.5 standard deviations in CSF and serum and was common in multiple populations. IL-34 deficiency reshaped CSF proteomic networks, downregulating axon guidance and microglial support modules while upregulating inflammatory and extracellular matrix signatures, and showed pleiotropic associations with neurological, inflammatory, and metabolic traits. Transcriptomic analysis of sorted microglia from healthy 9-month-old IL-34KO compare to wild-type mice revealed a profound pro-inflammatory and disease-associated microglial transcriptional program enriched for disease-associated microglia (DAM) signatures, inflammatory pathways, and AD risk genes including *APOE, CLU,* and *CASS4*. In APP/PS1 mice, genetic IL-34 deletion selectively depleted homeostatic gray-matter microglia, disrupted microglial tiling, and impaired plaque encapsulation, resulting in altered amyloid structure and enhancing neuritic injury. Concordantly, AD patients homozygous for IL-34-Y213X displayed markedly reduced cortical microglial density and increased microglial spatial dispersion, indicating a breakdown of the microglial network organization in the human brain.

**Interpretation:** A common human IL-34 LOF variant creates a naturally occurring model of IL-34 deficiency that links microglial survival, CSF network signatures, and amyloid pathology in both mice and humans. Importantly, IL-34 deficiency alone is sufficient to induce inflammatory, AD-associated microglial states beyond simply reducing microglial number. These findings identify IL-34/CSF1R signaling as a critical determinant of microglial resilience and a potential upstream pathway linking human genetic variation to AD susceptibility, highlighting IL-34-dependent pathways as promising targets for disease modification.

**Funding:** This work was supported by grants from the Spanish Ministerio de Ciencia, Innovación y Universidades/FEDER/UE (PID2024-157400OB-I00) and FORTALECE program (FORT23/00008; Instituto de Salud Carlos III, Spain) to RRL and JLV, ISCIII of Spain co-financed by FEDER funds (European Union) through grants PI24/00308 (JV) and CIBERNED collaborative grant 2022/01 to JV, PID2023-147125OB-I00 and CEX2023-001386-S (Severo Ochoa Programme) to SMTBC. A.R. is supported by STAR Award. University of Texas System. Tx, United States, The South Texas ADRC. National Institute of Aging. National Institutes of Health. USA. (P30AG066546), the Keith M. Orme and Pat Vigeon Orme Endowed Chair in Alzheimer’s and Neurodegenerative Diseases (2024-2025) and Patricia Ruth Frederick Distinguished Chair for Precision Therapeutics in Alzheimer’s and Neurodegenerative Diseases (2025-2028). AR is also supported by the Agency for Innovation and Entrepreneurship (VLAIO) grant N° PR067/21 for the HARPONE project and the ADAPTED project the EU/EFPIA Innovative Medicines Initiative Joint Undertaking Grant N° 115975 and CIBERNED (ISCIII).

## Introduction

Alzheimer’s disease (AD) is the leading cause of dementia worldwide and represents one of the greatest public health challenges of nowadays(*1*). Despite decades of research, effective disease-modifying therapies remain limited, largely because the cellular and molecular routes that drive pathogenesis, and define therapeutic windows, are still incompletely defined(*2, 3*). Human genetics has reshaped this landscape by consistently implicating innate immune pathways in AD susceptibility, placing microglia at the center of causal biology and motivating efforts to pinpoint the specific immune mechanisms that modify disease risk and progression(*4–9*). These findings underscore the urgent need to dissect the immune-related mechanisms and to identify therapeutic targets that can be leveraged to alter disease course.

Microglia, the resident immune cells of the central nervous system (CNS), arise from yolk-sac progenitors, colonize the brain during embryogenesis, and persist as long-lived tissue macrophages with slow but continuous turnover throughout life, that support neuronal wiring, synaptic maintenance and neuroimmune surveillance throughout life(*10–15*). In AD, microglial actions are increasingly understood as stage- and context-dependent: early responses can be protective, supporting tissue homeostasis and limiting damage, whereas failure of homeostatic programs and sustained activation may become maladaptive, contributing to synaptic dysfunction and neurodegeneration(*10–12, 14, 15*). This duality makes it essential to identify molecular pathways that maintain microglial fitness and homeostasis in vulnerable brain regions, and to define how inherited perturbations of those pathways translate into human disease.

A core trophic axis for microglial survival and proliferation is mediated by colony-stimulating factor 1 receptor (CSF1R), which is activated by colony-stimulating factor 1 (CSF1) and by the high-affinity ligand interleukin-34 (IL-34)(*16–18*). Although both ligands converge on CSF1R signaling, IL-34 exhibits distinct spatial and temporal regulation that is particularly relevant for long-lived microglial populations in the cortex and hippocampus, regions prominently affected in AD(*16, 17*). In adult brain, a large fraction of gray-matter microglia depend on IL-34 for survival, and IL-34 deficiency leads to marked reductions of homeostatic microglia, especially in cortex and hippocampus, underscoring a non-redundant role for IL-34 in sustaining microglial homeostasis and CNS integrity(*16–18*). These observations position IL-34 not simply as a generic growth factor, but as a regionally tuned regulator of microglial maintenance, precisely the type of biology that could shape susceptibility to neurodegeneration when genetically perturbed.

Supporting this hypothesis, convergent human genomic and proteomic studies have increasingly nominated IL-34 as a candidate AD locus. Marioni et al. first suggested IL-34 as a candidate signal in AD, and we subsequently identified a stop-gain mutation in IL-34 (p.Tyr213Ter, Y213X; rs4985556) associated with increased AD risk(*19*). This association was later validated in the largest AD meta-GWAS to date conducted by the European Alzheimer’s Disease Biobank(*20*). Extending genetic association to molecular consequence, our recent pQTL analysis of the global cerebrospinal fluid (CSF) proteome linked the same IL-34 variant to chemokine levels in CSF, implicating this AD genetic risk factor as a modulator of IL-34-dependent inflammatory networks within the CNS(*21*). Together, these data argue that IL-34 sits at the intersection of microglial homeostasis and human AD susceptibility, but they also highlight a critical gap: the systemic and CNS consequences of human IL-34 deficiency, including its biochemical signature, downstream proteomic networks, and neuropathological correlates, remain essentially undescribed.

Experimental studies further support the idea that IL-34-CSF1R signaling can shape AD-relevant phenotypes, while also emphasizing the need for human validation. In transgenic AD mouse models, intracerebroventricular IL-34 administration has been reported to ameliorate behavioral deficits, reduce oligomeric Aβ levels and promote microglial production of anti-inflammatory mediators such as TGF-β1, whereas IL-34 knockout (KO) mice show loss of homeostatic microglia, synaptic pathology and increased neuronal vulnerability in disease or injury contexts(*16, 18, 22*). More broadly, microglial proliferation and survival in AD are strongly influenced by CSF1R signaling, and proliferation levels correlate with disease severity in both humans and mice(*23, 24*). Given IL-34’s high affinity for CSF1R and its key role in sustaining microglia in cortex and hippocampus, inherited IL-34 loss-of-function (LOF) provides a plausible mechanistic bridge from genetic risk to impaired microglial homeostasis in the very regions most vulnerable in AD(*20*). Yet, despite this collective evidence, the phenotypic consequences of IL-34 deficiency in humans-including its biochemical signature, downstream proteomic networks, and neuropathological correlates-remain essentially undescribed.

Here we present a comprehensive characterization of IL-34 deficiency caused by the Y213X variant in the context of AD and validate key findings in an established model of the disorder. Specifically, we (i) demonstrate loss of human IL-34 protein in plasma and CSF of Y213X mutation carriers, (ii) identify CNS proteome networks associated with IL-34-dependent biology in humans, (iii) describe brain pathological features in naturally occurring homozygous carriers of the Y213X mutation, and (iv) generate and characterize IL-34KO mice, including their cross with APP/PS1 transgenic AD model. Together, these studies define IL-34 deficiency due to Y213X as a human immunogenetic entity, connect a specific LOF allele to molecular and neuropathological consequences, and establish a framework for translational strategies aimed at restoring IL-34-CSF1R signaling in individuals with deficient levels.

## Results

### IL-34-Y213X is a Common LOF Variant that Causes Profound IL-34 Deficiency in Humans

To investigate the functional impact of the IL-34-Y213X (rs4985556) nonsense mutation in humans, we quantified IL-34 levels in paired CSF and serum samples collected under standardized fasting conditions. Across both biofluids, we observed a clear, dose-dependent effect of genotype on cytokine levels. Heterozygous carriers (Y213X/+) showed markedly reduced IL-34 compared to wild-type alleles carriers, whereas homozygous carriers (Y213X/Y213X) exhibited near-complete loss of detectable IL-34. These effects were large in magnitude and highly significant, consistent with IL-34-Y213X acting as a strong LOF allele that generates a bona fide *in vivo* model of IL-34 deficiency in humans (Figure 1A, B, Supplementary Table 1).

**Figure 1:**
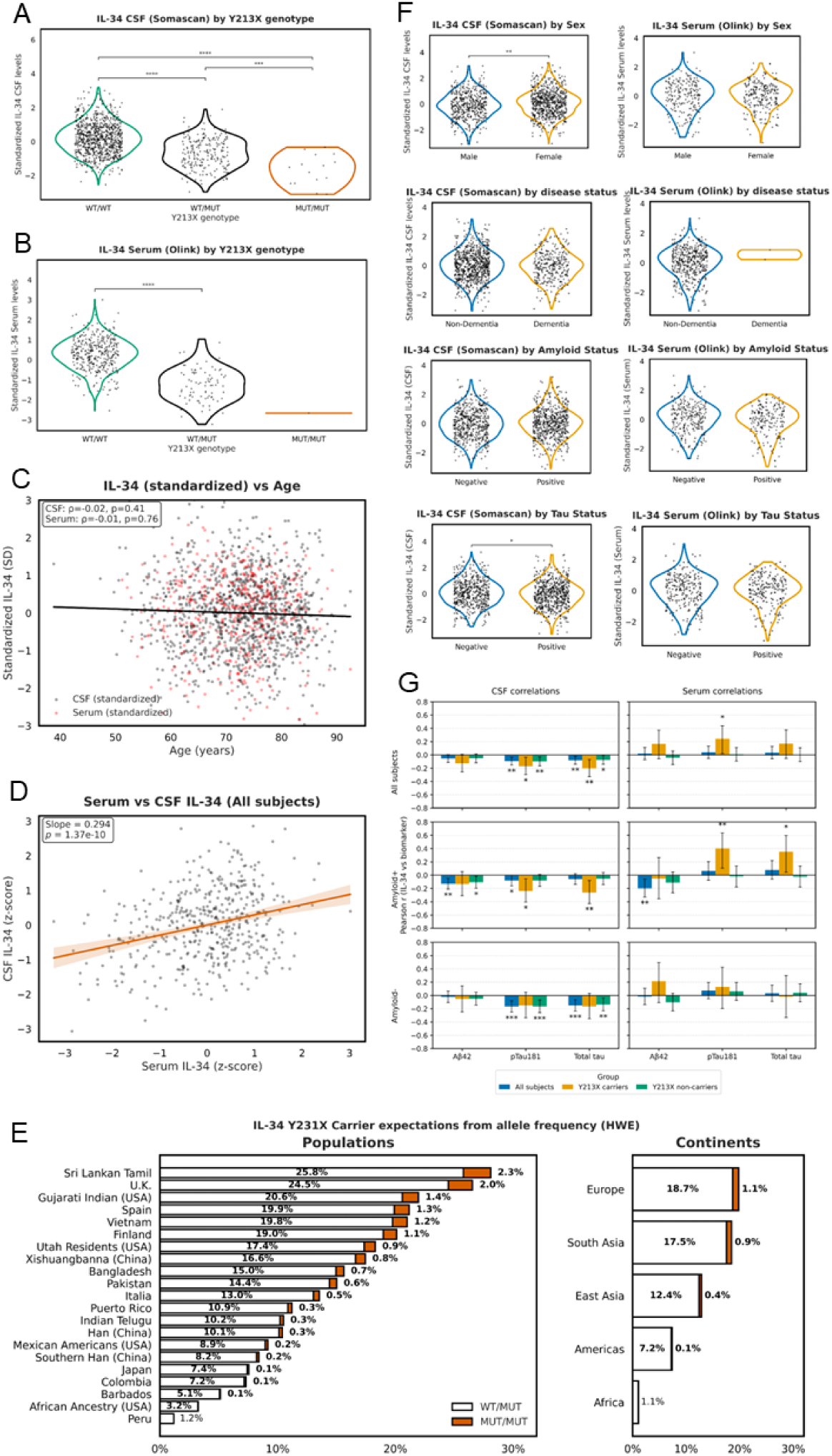
IL-34 levels in CSF and serum by Y213X genotype and clinical context. (A) Standardized IL-34 levels measured in cerebrospinal fluid (CSF; SomaScan) across Y213X genotypes (WT/WT, WT/MUT, MUT/MUT). (B) Standardized IL-34 levels measured in serum (Olink) across Y213X genotypes. (C) Relationship between standardized IL-34 levels and age, shown for CSF (gray) and serum (red). (D) Correlation between serum and CSF IL-34 levels across all subjects. (E) Expected Y213X carrier and homozygote frequencies across populations and continents derived from allele frequency under Hardy–Weinberg equilibrium. (F) IL-34 levels in CSF and serum stratified by sex, dementia status, amyloid status, and tau status. (G) Pearson correlations between IL-34 and CSF Alzheimer’s disease biomarkers (Aβ_42_, pTau181, and total tau), shown for all subjects and after stratification by Y213X carrier status and amyloid status. Error bars denote 95% confidence intervals. Asterisks indicate significance thresholds (∗ p < 0.01, ∗∗ p < 0.001).

Importantly, the impact of the IL-34-Y213X variant on CSF IL-34 levels was replicated across three independent experiments using two orthogonal proteomic platforms (two SomaScan experiments and one Olink immunoassay; Supplementary Figure 1). This cross-platform concordance reduces the likelihood of assay-specific artifacts and supports the conclusion that Y213X carriers experience a true biological deficit in IL-34 rather than spurious measurement variation.

To determine whether IL-34 protein levels were primarily driven by genetic or non-genetic factors, we fitted multivariable linear models including age at lumbar puncture, sex, dementia status, and ATN biomarker positivity. Demographic and clinical variables collectively explained minimal variance in IL-34 levels across both platforms (CSF SomaScan: R² = 0.018, n = 1,130; serum Olink: R² = 0.011, n = 460). In contrast, IL-34-Y213X dosage alone accounted for 16.5% of variance in CSF (R² = 0.165, n = 1,138) and 37.4% in serum (R² = 0.374, n = 461). Adding genotype to the demographic/clinical model yielded substantial incremental gains (CSF: ΔR² = 0.163, full model R² = 0.181, p = 3.7 × 10^−46^; serum: ΔR² = 0.364, full model R² = 0.375, p = 5.1 × 10^−47^), confirming that IL-34-Y213X is the dominant determinant of IL-34 protein abundance in this cohort. These findings establish a genetically anchored protein phenotype that can be precisely quantified across biofluids and is directly relevant to IL-34-mediated AD risk (Figure 1A, B, F).

To place these biochemical findings in a broader clinical context, we next interrogated large-scale phenome-wide association resources (UK Biobank, FinnGen and the GWAS Catalog)(*25, 26*) for traits associated with IL-34-Y213X. Beyond its impact on IL-34 protein levels itself, the mutation showed pleiotropic associations with a range of neurological, inflammatory and metabolic traits, indicating that IL-34 deficiency may have systemic consequences that extend beyond AD (Supplementary Tables 2 and 3). Leveraging data extracted from the GWAS Catalog(*27*), we identified independent variants near or within the human IL-34 locus that modulate IL-34 protein expression and are associated with additional phenotypes, further supporting the view that IL-34 biology is embedded within a broader regulatory and pleiotropic landscape (Supplementary Tables 4 and 5; Supplementary Results).

To assess the worldwide distribution of the IL-34-Y213X (rs4985556, p.Tyr213Ter) nonsense mutation, we examined allele frequencies across the 26 populations of the 1000 Genomes Project Phase 3 (n = 2,504 individuals; 5,008 chromosomes). Data were obtained from LDlink (LDpop module) and aggregated into continental super-populations (AFR, AMR, EAS, EUR, SAS) (Figure 1E, Supplementary table 6).

As shown in Figure 1E (left), the IL-34-Y213X mutation is geographically structured, with striking differences across populations. The highest allele frequency was observed in the Sri Lankan Tamil in the UK (STU, South Asian ancestry; 15.2%; 31/204 chromosomes), followed by British in England and Scotland (GBR, 14.3%; 26/182 chromosomes) and Kinh in Vietnam (KHV, 11.1%; 22/198 chromosomes). Other European (IBS, Iberian from Spain, 11.2%) and East Asian (CHB, Han Chinese from Beijing, 5.3%) populations also carried the variant at appreciable frequencies. In contrast, the variant was absent from multiple African cohorts (YRI, LWK, GWD, MSL, ESN) and extremely rare in admixed American populations, with the lowest non-African frequency observed in Peruvians from Lima (PEL, 0.59%; 1/170 chromosomes).

When aggregated at the continental level, the European (EUR) and South Asian (SAS) super-populations carried the greatest burden of the Y213X allele, with minor allele frequencies (MAF) of 10.4% and 9.7%, respectively. Under Hardy–Weinberg equilibrium, these values translate into 18–20% of individuals predicted to be heterozygous carriers (haploinsufficient) and ∼1% homozygous (IL-34 full deficiency). East Asian populations (EAS) showed an intermediate frequency (MAF = 6.7%), corresponding to ∼12% heterozygotes and 0.4% homozygotes. In contrast, the American (AMR) populations carried the mutation at lower levels (MAF = 3.7%; ∼7% heterozygotes, 0.1% homozygotes), while Africans (AFR) were virtually devoid of the mutation (MAF = 0.5%; haploinsufficiency ∼1%, no homozygotes observed) (Figure 1E, right). Together, these findings indicate that IL-34 deficiency is not a rare genetic condition but instead represents a geographically structured trait. The highest prevalence is found in South Asians and Europeans, where up to one in five individuals may be haploinsufficient and ∼1 in 100 may completely lack IL-34 due to homozygosity for Y213X, whereas African populations are essentially spared from this mutation.

Collectively, our results establish IL-34-Y213X as a common, high-impact LOF variant that produces profound IL-34 protein level deficiency in humans and is associated with a broader pleiotropic burden. This naturally occurring “human IL-34 knockout” provides a strong entry point for dissecting how IL-34 signaling shapes microglial biology, CSF proteomic networks, and vulnerability to AD in the subsequent sections.

### Human Serum and CSF IL-34 levels partially correlate and diverge in their associations with AD biomarkers depending on Y213X mutation and amyloid status

Next, we examined in greater detail the relationship between IL-34 and AD biomarkers across central and peripheral compartments (Figure 1G). CSF and serum IL-34 levels were moderately correlated (β = 0.29, p = 1.37×10⁻¹⁰), suggesting limited concordance between central and peripheral IL-34 levels. Within CSF, IL-34 showed consistent inverse associations with tau-related pathology, even after adjusting for age, sex and MMSE. The largest effect sizes were observed in Y213X carriers, where CSF IL-34 correlated with total tau (r = –0.20, 95% CI [–0.33, –0.07], p = 0.0029) and pTau181 (r = –0.17, 95% CI [–0.30, – 0.04], p = 0.012). In amyloid-positive subjects carrying the Y213X mutation, these effects were even more pronounced (pTau181: r = –0.24, 95% CI [–0.41, –0.06], p = 0.010; total tau: r = –0.26, 95% CI [–0.43, –0.08], p = 0.0048). Associations were also present in amyloid-negative groups, most notably for pTau181 and total tau in non-carriers Y213X (r ≈ –0.16 to – 0.17).

Although only a subset of these correlations survived Bonferroni correction (supplementary table 7, 8), the consistently negative direction and overlapping confidence intervals across subgroups reinforce the interpretation that lower CSF IL-34 is linked to greater tau-related neuronal injury. Importantly, effect sizes rather than corrected p-values may provide a more realistic picture here, given the marked imbalance in sample sizes (e.g. ∼4× more non-carriers than carriers), which strongly impacts statistical power and the probability of detecting true effects in smaller subgroups.

By contrast, serum IL-34 did not reproduce these CSF associations (Figure 1G). Associations with tau-related markers were weaker and often reversed in sign, particularly in amyloid-positive carriers (pTau181: r = +0.17, 95% CI [–0.05, +0.36], p = 0.12; total tau: r = +0.29, 95% CI [–0.01, +0.53], p = 0.056). Although suggestive of a potential peripheral immune signature, these findings did not reach statistical significance and should be considered with caution and require validation in larger cohorts. Together, these results indicate that CSF IL-34 more directly reflects AD-related processes in a mutation- and amyloid-dependent manner, whereas serum IL-34 may instead capture a weaker, systemic response with less clear relevance to tau pathology in the brain (Supplementary table 7).

### Convergence of Correlation Networks and Genetic Deficiency of the Human IL-34 Cytokine in Cerebrospinal Fluid

Having established that IL-34-Y213X produces a profound, genetically anchored deficiency of IL-34 in humans, we next explored how variation in IL-34 levels is embedded in broader CSF protein networks. To investigate the functional impact of IL-34 deficiency on the human CSF proteome, we combined correlation network analysis of IL-34 levels across the cohort with differential expression profiling in *IL-34* Y213X homozygotes compared to matched wild-type controls. Both approaches revealed highly interconnected protein modules and pointed to a consistent biological signature linking IL-34 deficiency to alterations in synaptic guidance, microglial function, and extracellular regulation (Figures 2A-C and 3A-D).

**Figure 2:**
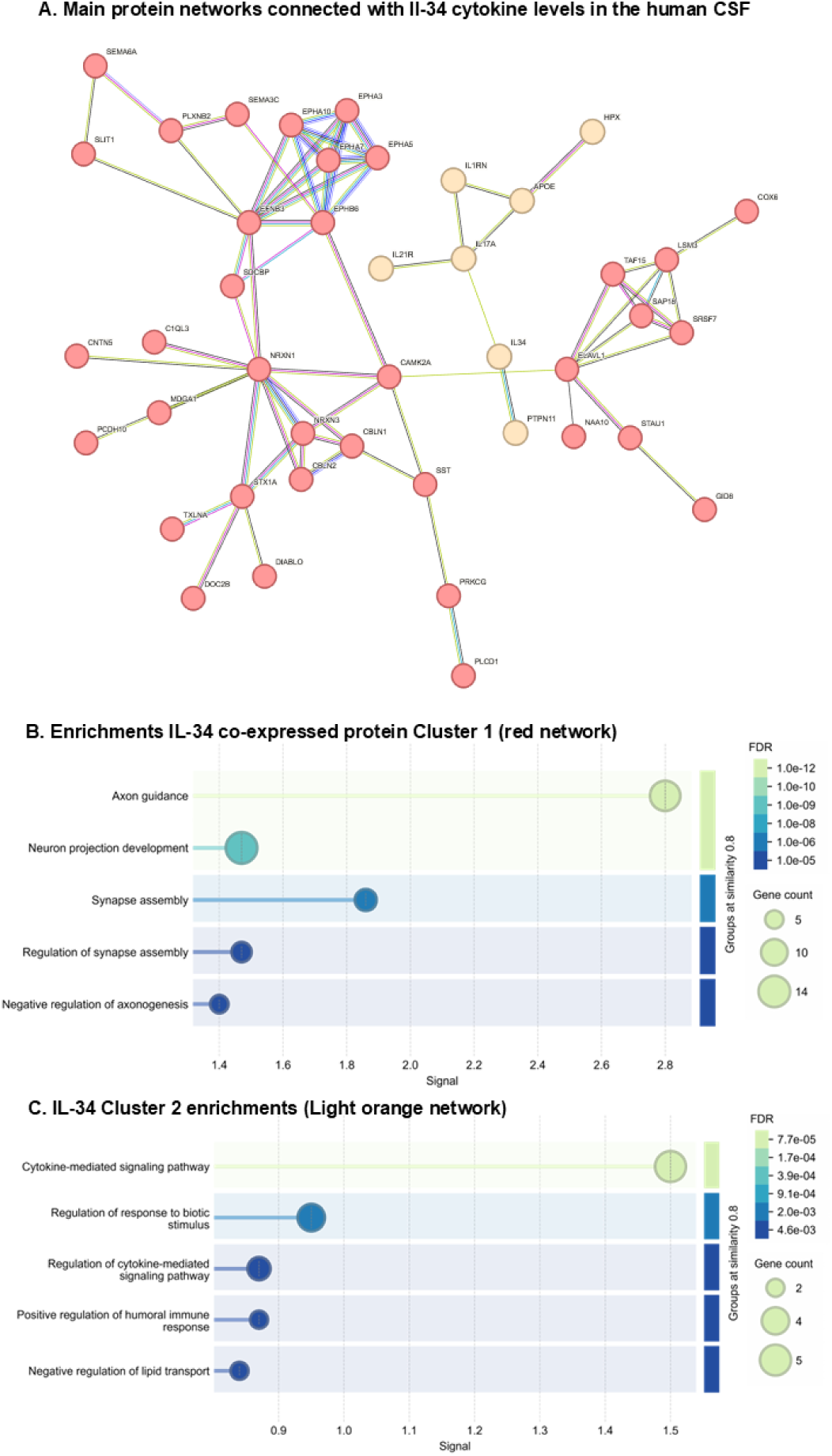
Protein interaction networks associated with IL-34 in human CSF. (A) STRING protein–protein interaction network of proteins associated with CSF IL-34 levels, highlighting two major modules. (B) Gene Ontology enrichment results for the module positively associated with IL-34 (red network), showing enrichment for neuronal and synaptic biology. (C) Gene Ontology enrichment results for the module negatively associated with IL-34 (light orange network), enriched for cytokine-mediated signaling and related immune processes. Bubble size reflects gene counts and color reflects FDR-adjusted significance.

**Figure 3:**
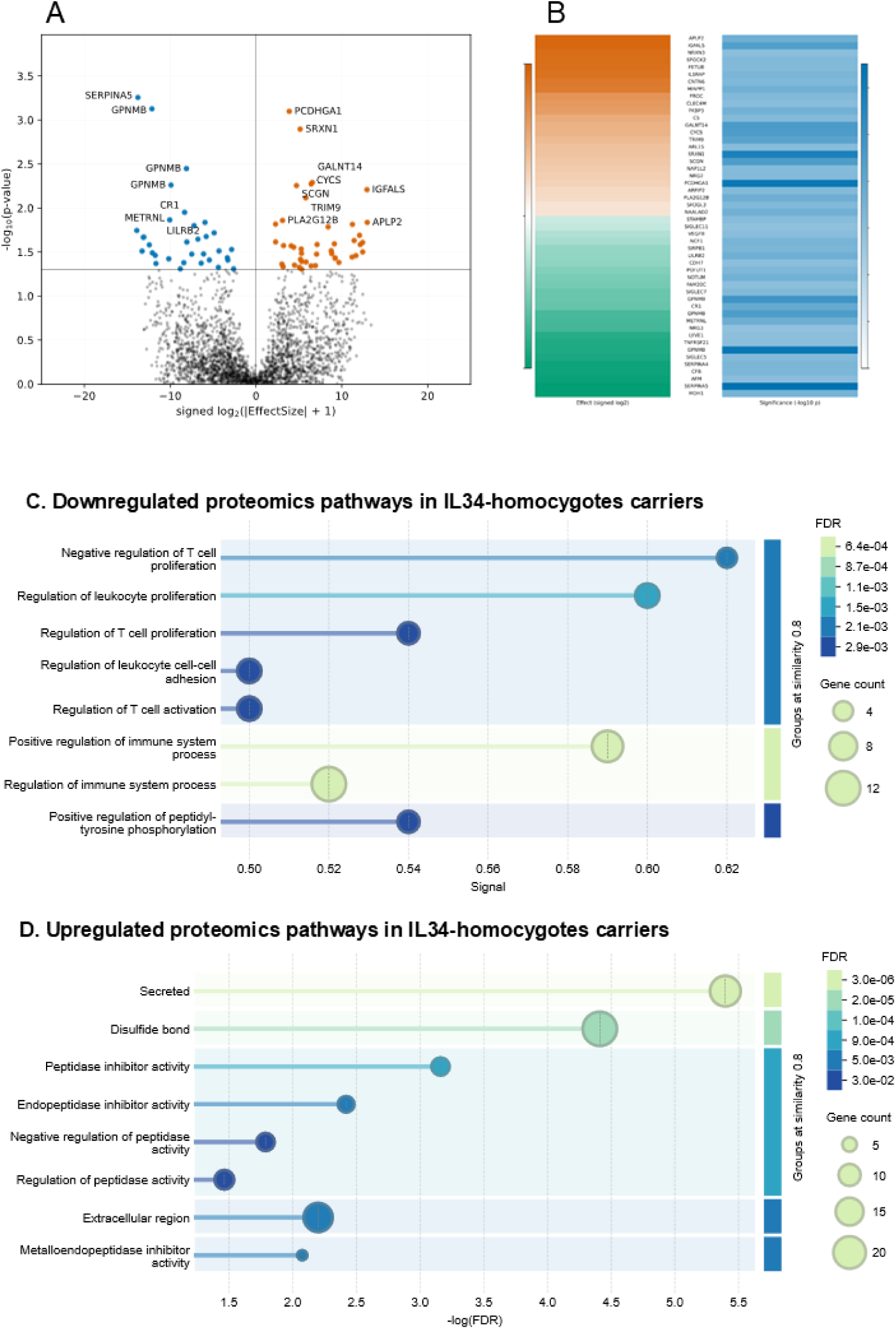
Differential CSF proteomic signatures in IL-34 Y213X homozygotes. (A) Volcano plot comparing CSF proteomes from IL-34 Y213X homozygotes and matched controls, highlighting nominally significant proteins. (B) Heatmap summarizing effect sizes and statistical significance for the selected proteins shown in panel A. (C) Functional enrichment of pathways represented among proteins decreased in IL-34 homozygotes. (D) Functional enrichment of pathways represented among proteins increased in IL-34 homozygotes. Bubble size reflects gene counts and color denotes FDR-adjusted significance.

Specifically, the correlation analysis of IL-34 cytokine levels in the CSF with the global SomaScan proteome identified 98 proteins significantly co-regulated with IL-34 (|r| > 0.2, p< 6.05E-14) (Figure 2). Of these, 63 proteins were positively correlated with IL-34 and therefore downregulated when IL-34 levels were low (Supplementary Figure 3), while 37 proteins were negatively correlated and thus upregulated in the setting of IL-34 deficiency (Supplementary Figure 3). The global STRING PPI map (Figure 2A) displayed strong connectivity (PPI enrichment p < 1.75 × 10⁻^9^), which, when partitioned by correlation directionality (Supplementary Figure 3), resolved into different biologically distinct clusters.

Cluster 1, composed of proteins positively correlated with IL-34 levels, was strongly enriched for axon guidance (GO:0007411; 28/63 proteins; FDR = 2.1 × 10⁻⁶), synapse organization (GO:0050808; 16/63; FDR = 8.3 × 10⁻⁵), and CSF1R signaling in myeloid cells (REAC:R-HSA-6785807; 12/63; FDR = 2.9 × 10⁻³). Among the strongest associations in this network were NRXN1 (r = 0.28, p = 1.2 × 10⁻²⁶), EPHA5 (r = 0.27, p = 2.5 × 10⁻²⁵), and CAMK2A (r = 0.27, p = 3.1 × 10⁻²⁵), which also acted as hubs with the highest degree of connectivity. These findings highlight IL-34’s role in sustaining axon projection pathways, synaptic adhesion, and microglial–neuronal communication. In contrast, Cluster 2, mostly composed of proteins negatively correlated with IL-34 and therefore upregulated when IL-34 was low, was enriched for RNA splicing (GO:0008380; 10/37 proteins; FDR = 4.1 × 10⁻⁵), interleukin-1 signaling (REAC:R-HSA-9020702; 6/37; FDR = 2.8 × 10⁻³), collagen trimerization and extracellular matrix assembly (GO:0032963; 7/37; FDR = 9.6 × 10⁻³). Key drivers of this module included PLXNB2 (r = –0.27, p = 1.4 × 10⁻²⁴), CBLN1 (r = –0.27, p = 2.6 × 10⁻²⁴), MDGA1 (r = –0.26, p = 5.7 × 10⁻²²), and IL21R (r = –0.25, p = 1.0 × 10⁻²⁰), pointing to a coordinated upregulation of RNA-binding proteins, synaptic adhesion complexes, pro-inflammatory cytokine receptors, and extracellular remodeling pathways in the context of reduced levels of IL-34 in human CSF. Thus, Figure 2 illustrates the global network structure, while Supplementary Figure 2 disentangles the directionality of the associations, clearly separating proteins downregulated *versus* upregulated with low IL-34.

To test whether genetic deficiency of IL-34 recapitulates these network-derived associations, we next examined CSF proteomes exclusively from IL-34 Y213X homozygotes (n=17) and matched wild-type controls (see methods). Because only seventeen homozygous carriers were available, the statistical power of this experiment was limited. Accordingly, we focused on the subset of proteins showing nominal statistical significance (p < 0.05) and the most consistent effect sizes.

STRING analysis of these nominally significant proteins confirmed that they assemble into functionally coherent networks (especially for downregulated proteins with PPI enrichment (p < 6.24 × 10^−8^). The downregulated module was enriched for processes suppressing the immune system activation, including regulation of immune system process (GO:0002684; FDR = 6.4 × 10⁻⁴), regulation of leukocyte proliferation (GO:0070663; FDR = 1.7 × 10⁻³), and T cell activation (GO:0042110; FDR = 3.0 × 10⁻³). The upregulated module showed enrichments in extracellular biology, with strongest signals for endopeptidase inhibitor activity (GO:0004866; FDR = 3.0 × 10⁻⁶), regulation of peptidase activity (GO:0052547; FDR = 2.0 × 10⁻⁵), and extracellular region localization (GO:0005576; FDR = 9.0 × 10⁻⁴). Thus, even under reduced sample size, genetic IL-34 deficiency reproducibly mirrored the network-level predictions of correlation analyses, demonstrating both a collapse of microglial-immune-extracellular signaling modules and a compensatory induction of protease inhibition and stress-response proteins.

The volcano plot in Figure 3A-B highlights these top signals. Downregulated proteins included GPNMB and CR1, both key mediators of microglial activation and complement regulation, together with other glycosaminoglycan, and heparin-binding proteins. This pattern is consistent with reduced microglial signaling capacity and impaired extracellular guidance when IL-34 is genetically disrupted. In contrast, upregulated proteins included members of the SERPIN family of protease inhibitors, as well as PCDHGA1, SRXN1, CYCS, and GALNT14, reflecting a compensatory increase in protease inhibition, alterations in mitochondrial metabolism, and changes in extracellular plasticity.

Together, the correlation networks (Figure 2 and Supplementary Figure 2) and the differential expression analysis (Figure 3) identified a consistent set of IL-34–associated changes in the human CSF proteome. Lower IL-34 levels were associated with reduced abundance of axon guidance and microglia-related proteins, including NRXN1, EPHA5, GPNMB and CR1, and with increased abundance of RNA-binding proteins, synaptic adhesion molecules (CBLN1, MDGA1), cytokine receptors (IL21R) and extracellular protease inhibitors (SERPIN family members), consistent with prior works (*10, 16–18, 28, 29*).

### IL-34 deficiency induces a profound pro-inflammatory microglial phenotype and promotes a transcriptional shift toward AD risk-associated programs

Having thoroughly characterized the impact of the IL-34-Y213X mutation in humans, we next sought to establish an AD animal model lacking IL-34. The rationale for using a KO mouse model to investigate the effects of the human Y213X mutation was to mimic the profound IL-34 deficiency observed in human IL-34-Y213X carriers (Figure 1). As a first step, we analyzed microglial density in different brain regions (cortex and hippocampus) in adult IL-34KO animals. Our analysis revealed a marked reduction in the number of Iba1^+^ homeostatic microglial cells in the cortex (Figure 4A-B), and a similar decrease in the hippocampus (Figure 4A, C), consistent with previous reports (*30*).

**Figure 4:**
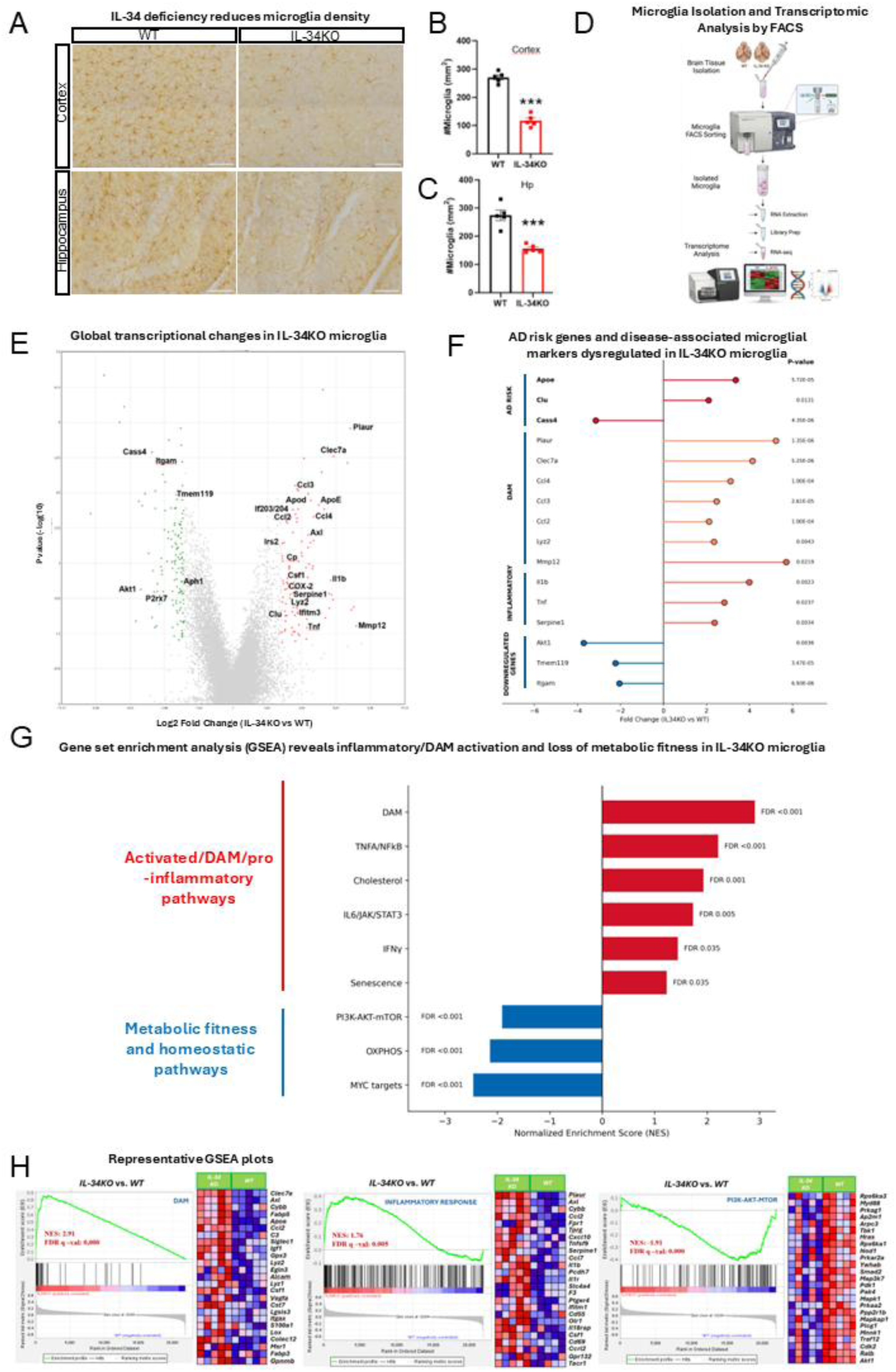
Loss of IL-34 reduces microglial density and promotes a disease-associated, pro-inflammatory transcriptional program. (A) Representative Iba1 immunostaining in cortex and hippocampus of WT and IL-34KO mice. Scale bar: 100 µm. (B-C) Quantification of microglial density (Iba1⁺ cells/mm²) in cortex (B) and hippocampus (C). (D) Schematic overview of microglia isolation by FACS and downstream transcriptomic analysis. (E) Volcano plot displaying differentially expressed genes between WT and IL-34KO microglia. Selected upregulated (red) and downregulated (green) genes are indicated. (F)Differential expression of Alzheimer’s disease (AD) risk genes, disease-associated microglia (DAM) markers, inflammatory mediators, and homeostatic microglial genes in IL-34KO microglia. Values represent log₂ fold change (IL-34KO vs. WT). Corresponding p-values are shown on the right. (G) Gene Set Enrichment Analysis (GSEA) demonstrating enrichment of DAM and pro-inflammatory pathways, including TNFA/NFκB signaling, IL6/JAK/STAT3 signaling, interferon-γ response, cholesterol metabolism, and senescence pathways, together with impairment of pathways associated with metabolic fitness and microglial homeostasis, including PI3K-AKT-mTOR signaling, oxidative phosphorylation (OXPHOS), and MYC targets. Bars represent normalized enrichment scores (NES). FDR values shown correspond to GSEA false discovery rate-adjusted q-values (FDR q-values). (H) Representative GSEA enrichment plots and associated heatmaps illustrating activation of DAM and inflammatory-response gene signatures and impairment of PI3K-AKT-mTOR signaling in IL-34KO microglia. NES, normalized enrichment score; FDR q-value, false discovery rate-adjusted q-value. Data in B-C are shown as mean ± SEM; asterisks indicate statistical significance (***p < 0.001).

To investigate the role of IL-34 in the maintenance of microglial homeostasis under physiological conditions, we performed transcriptomic profiling of FACS-isolated microglia from 9-month-old WT and IL-34KO mice (Figure 4D-F and Supplementary Table 9-11). Despite the absence of overt neurodegenerative pathology, IL-34 deficiency induced a profound transcriptional reprogramming of microglia, as illustrated by the marked segregation of differentially expressed genes in the volcano plot (Figure 4E). Differential expression analysis demonstrated robust induction of genes associated with inflammatory activation and disease-associated microglia (DAM), including *Clec7a, Apoe, Axl, Ccl2, Ccl3, Ccl4, Lyz2, Ifitm3, Plaur,* and *Mmp12* (Figure 4E-F). Consistent with activation of innate immune pathways, IL-34KO microglia also exhibited increased expression of pro-inflammatory mediators such as Il1b, Tnf, and Ptgs2/COX-2. Notably, several established AD genetic risk genes were significantly dysregulated, including increased expression of *Apoe* and *Clu* together with reduced expression of *Cass4* (Figure 4F). These transcriptional changes emerged in healthy 9-month-old mice lacking overt amyloid pathology or neurodegeneration, indicating that IL-34 deficiency alone is sufficient to perturb AD-relevant microglial programs. In parallel, IL-34KO microglia displayed reduced expression of genes associated with homeostatic microglial identity, including *Tmem119, Itgam, P2rx7*, and *Akt1*, consistent with a loss of canonical microglial homeostasis (Figure 4E-F).

To define the biological pathways underlying these transcriptional changes, we performed gene set enrichment analysis (GSEA). IL-34KO microglia showed significant enrichment of inflammatory and disease-associated pathways, including TNFα signaling via NFκB, inflammatory response, IL6/JAK/STAT3 signaling, interferon response and cholesterol (Figure 4G). Importantly, the strongest enrichment corresponded to a DAM gene signature (NES=2.9, FDR<0.001) (Figure 4G-H), demonstrating that IL-34 deficiency induces a coordinated neurodegeneration-associated microglial program rather than isolated inflammatory gene changes. In addition, IL-34KO microglia exhibited enrichment of senescence-associated pathways, a feature associated to AD pathology (*31*).

In contrast, pathways associated with metabolic fitness and cellular maintenance were significantly reduced in IL-34KO microglia. These included PI3K/AKT/mTOR signaling and oxidative phosphorylation, together with MYC target programs involved in cellular growth and maintenance (Fig. 4G-H). Collectively, these findings indicate that IL-34 deficiency does not merely reduce microglial abundance but drives the emergence of chronically activated, disease-associated microglia characterized by inflammatory activation, senescence-associated features, dysregulation of AD genetic risk pathways, and impaired metabolic fitness.

### Cross-species comparison supports partial concordance between human genetic IL-34 deficiency and IL-34KO microglia

We next asked whether the proteomic signature of genetic IL-34 deficiency in humans aligned with the transcriptional response observed in IL-34KO mouse microglia. This comparison is deliberately stringent, because it integrates a global human CSF proteomic readout with a purified mouse microglial transcriptomic dataset. We compared the CSF SomaScan differential signature from IL-34 Y213X homozygotes versus matched WT alleles carriers controls with the ranked IL-34KO versus WT mouse microglial transcriptome (Supplementary Figure 4). After excluding IL-34 itself, 42 proteins were nominally elevated and 30 reduced in human IL-34-Y213X homozygotes relative to matched controls; following ortholog mapping, these defined the Y213X-up (n = 42) and Y213X-down (n = 30) modules (Supplementary Figure 4).

The human Y213X-up module showed a positive enrichment trend among genes upregulated in IL-34KO microglia (NES = 1.29, P = 0.098, FDR = 0.197), whereas the Y213X-down module did not show significant enrichment in the opposite direction (NES = −0.79, P = 0.752, FDR = 0.752). Although categorical module enrichment did not reach FDR significance, rank-level comparison across all mapped genes revealed a modest but significant positive concordance between the human IL-34-deficiency proteomic signature and the mouse IL-34KO microglial transcriptome (Spearman rho = 0.085, P = 2.77 x 10^−4^; Pearson r = 0.082, P = 4.48 x 10^−4^). These findings indicate partial cross-species molecular alignment between human genetic IL-34 deficiency and experimental IL-34 loss in mouse microglia.

Inspection of the concordant signals highlighted several biologically coherent candidates (Supplementary Table 12). APLP2, SRXN1, FLT1 and MDGA1 were increased in both IL-34 Y213X homozygous human CSF proteomes and IL-34 knockout mouse microglia, implicating amyloid-related protein biology, redox-stress responses, vascular-inflammatory signaling and extracellular/synaptic adhesion remodeling as partially conserved consequences of IL-34 loss. Additional concordant upregulated signals included CYCS, GALNT14, NRG2 and NAALAD2, further supporting stress-response, glycosylation/extracellular plasticity and neuroglial remodeling axes. Conversely, TNFRSF21 and HCLS1 were reduced in both systems, suggesting that IL-34 deficiency also converges on selected immune-signaling and cytoskeletal programs. Importantly, this cross-species signal complements rather than replaces the mouse-specific AD-risk transcriptional program, in which IL-34KO microglia showed robust induction of DAM and inflammatory mediators, including Apoe, Clec7a, Axl, Ccl2, Ccl3, Ccl4, Plaur and Mmp12, together with increased Apoe and Clu and reduced Cass4. Thus, even across a highly orthogonal comparison of human CSF proteomics and mouse microglial transcriptomics, IL-34 deficiency converged on a subset of AD-relevant, stress, vascular-inflammatory and extracellular remodeling signals.

### IL-34 Deficiency Selectively Depletes Homeostatic Microglia and Reshapes Amyloid Pathology

We next crossed IL-34KO mice with APP/PS1 transgenic mice, and analyzed animals at 9-month-age, when APP/PS1 mice established amyloid pathology, altered microglial responses, and overt neuroinflammation(*32*). Our goal was to determine how IL-34 deficiency influences key features of amyloid pathology, including number and types of plaques, microglia clustering, and plaque toxicity. Total number and types of plaques were evaluated in terms of Aβ immunoreactivity and Methoxy-XO4 labeling, a marker of fibrillar amyloid-beta. Given the critical role of IL-34 in sustaining homeostatic microglia, and in light of growing evidence that homeostatic microglia are essential for the initial seeding and formation of amyloid plaques(*17, 18, 33–35*), we first examined the impact of IL-34 deficiency on amyloid plaque pathology. In APP/PS1 mice lacking IL-34, we observed a pronounced reduction in the survival of homeostatic (non-plaque-associated) microglia, with a ∼67% decrease compared to APP/PS1 controls (p < 0.001) (Figure 5I). Strikingly, analysis of total number of amyloid plaques revealed a pronounced reduction in IL-34KO-APP/PS1 mice (P = 0.002; Figure 5A, B and p = 0.049 in cortex and p = 0.029 in hippocampus; Figure 5D-F), underscoring the indispensable role of homeostatic microglia in the earliest stages of amyloid plaque initiation and growth(*36*).

**Figure 5:**
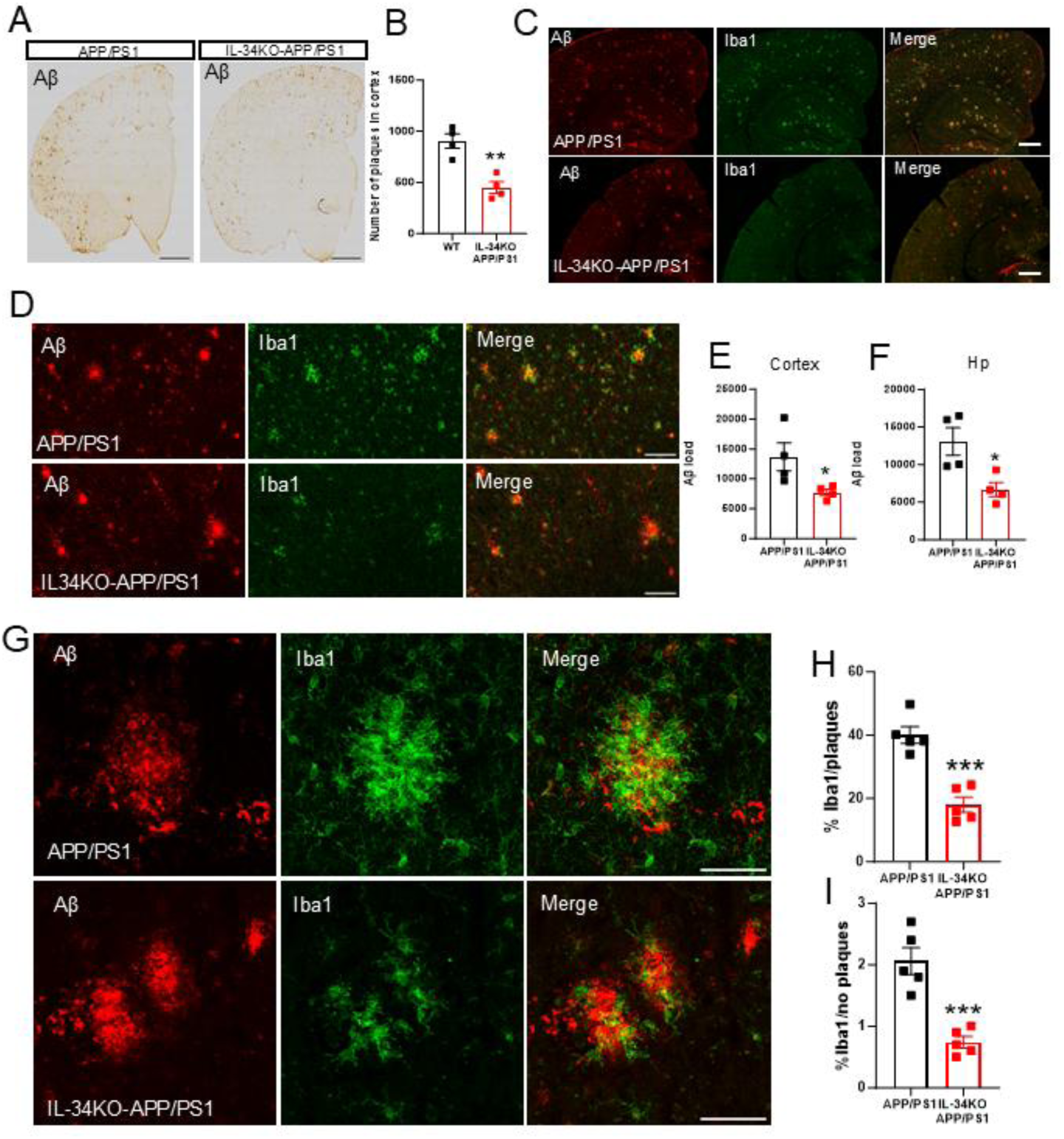
(A) Representative Aβ (BAM10) immunostaining in coronal sections from APP/PS1 and IL-34KO-APP/PS1 mice. Scale bar: 1 mm. (B) Quantification of cortical plaque number. (C, D) Low-magnification confocal images showing Aβ (red) and microglia (Iba1, green) in cortex of APP/PS1 and IL-34KO–APP/PS1 mice. Scale bar: 500 µm (E, F) Quantification of Aβ load in cortex (E) and hippocampus (F). (G) High-magnification confocal images illustrating microglial association with plaques. Scale bar: 50 µm. (H, I) Percentage of Iba1⁺ microglia per plaque (H) and proportion of microglia outside plaques (I). Data are shown as mean ± SEM; asterisks indicate statistical significance (*p < 0.05, **p < 0.01, ***p < 0.001).

Microglia normally cluster tightly around amyloid plaques, forming a physical barrier that promotes plaque compaction and reduces neurotoxicity(*37, 38*). Given the profound reduction in homeostatic microglia observed in IL-34KO animals, we next wondered whether this deficit affects the number of microglia available to engage and encapsulate plaques. IL-34 deficiency resulted in a dramatic reduction in microglial coverage of amyloid plaques compared with APP/PS1 controls (Figure 5G, H).

Deficiencies in TREM2 or its adaptor DAP12 are known to impair microglia clustering around plaques(*39–41*), a phenomenon attributed to their reduced ability to polarize processes toward the plaque surface and form an effective barrier(*41*). However, the phenotype observed under IL-34 deficiency differs fundamentally from that seen in TREM2- or DAP12-deficient microglia. In IL-34KO mice, microglia retained the capacity to polarize toward amyloid deposits but remained too few in number to achieve full plaque encapsulation, resulting in incomplete coverage of the plaque surface. This finding underscores that the sustained microglial response required for effective plaque clustering and maintenance of the microglial barrier is critically dependent on the IL-34/CSF1R proliferative axis.

Given the inability of IL-34KO animals to achieve effective plaque clustering, a plausible consequence would be an inadequate plaque compaction thus resulting in a non-uniform, irregular shape and a shift from compact to filamentous plaques. To test this possibility, we performed a quantitative analysis and found that small filamentous plaques were extremely dystrophic (Figure 6).

**Figure 6:**
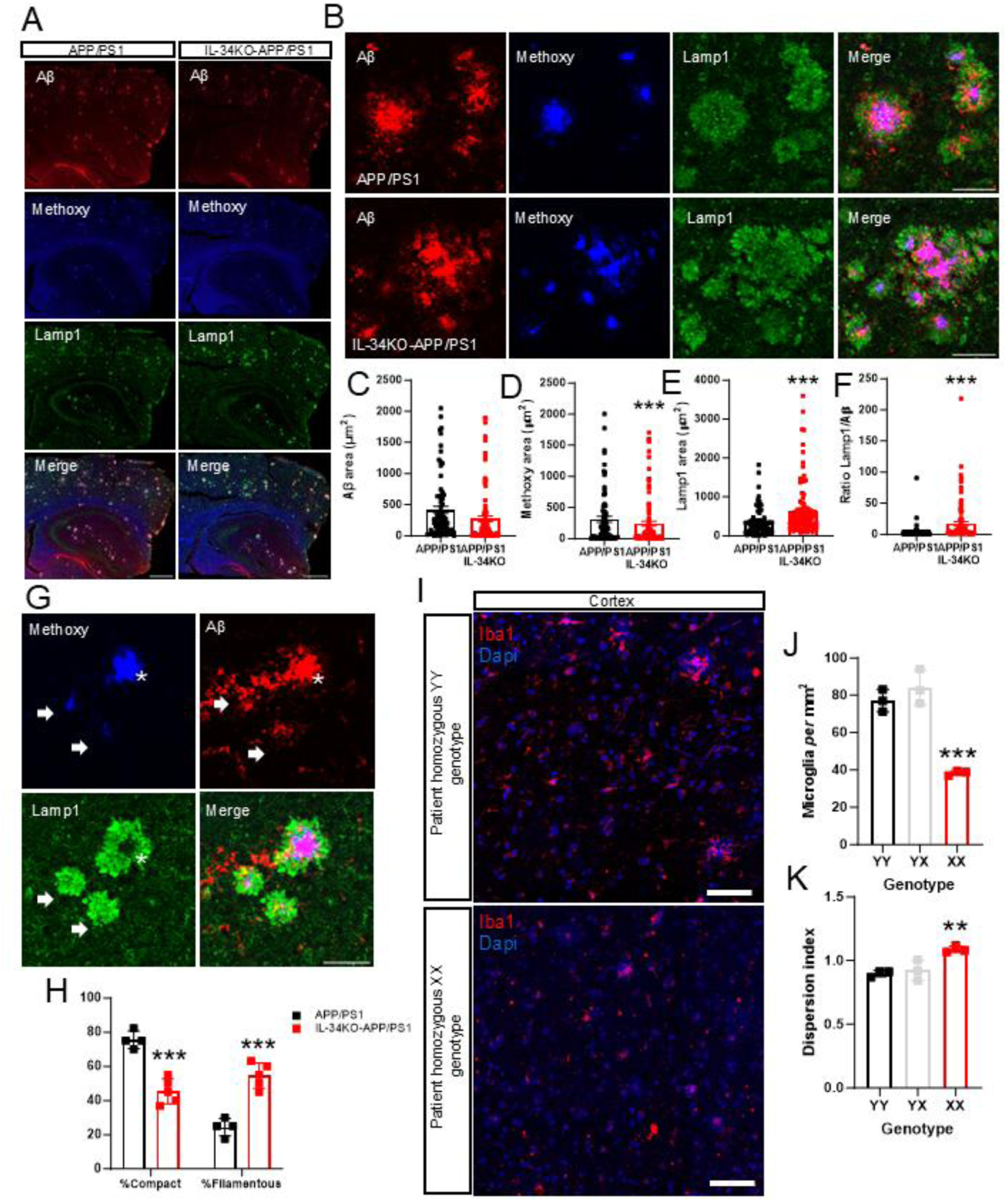
IL-34 deficiency alters plaque structure in APP/PS1 mice and is associated with reduced microglial density and altered spatial organization in human cortex. (A) Representative confocal images of Aβ (red), Methoxy-X04 (blue), and Lamp1 (green) in APP/PS1 and IL-34KO–APP/PS1 cortex. Scale bar: 500 µm. (B) Higher-magnification images showing Lamp1⁺ dystrophic neurites around plaques. Scale bar: 50 µm. (C-F) Quantification of Aβ plaque area (C), Methoxy-X04-positive compact core area (D), Lamp1⁺ dystrophic neurite area (E), and the Lamp1/Aβ ratio (F). (G) Example plaques illustrating compact (asterisk) and filamentous (arrows) morphologies. Scale bar: 50 µm. (H) Quantification of compact and filamentous plaque types. (I) Representative human cortical sections stained for Iba1 (microglia) with DAPI counterstain, shown for a YY homozygote and an XX homozygote. Scale bar: 100 µm. (J) Microglial density (cells/mm²) by Y213X genotype. (K) Dispersion index (spatial tiling metric) by Y213X genotype. Data are shown as mean ± SEM; asterisks indicate statistical significance (**p < 0.01, ***p < 0.001).

Together, these findings suggest IL-34 as a central regulator of homeostatic microglial maintenance and reveal that its absence profoundly reshapes the trajectory of amyloid pathology. By demonstrating that IL-34 deficiency drastically limits the pool of homeostatic microglia, disrupts microglial barrier formation despite preserved polarization capacity, reduces plaque number, and leads to poorly compacted, dystrophic amyloid deposits, our study uncovers an essential IL-34/CSF1R–dependent axis governing the earliest steps of plaque initiation, growth, and structural containment. These results position IL-34 as a previously unrecognized but indispensable determinant of microglial competence in AD and highlight its potential as a therapeutic target for modulating disease onset and progression.

### IL-34 Deficiency due to Y213X mutation Disrupts Microglial Homeostasis and Tiling in Human AD Cortex

Given the profound impact of IL-34 deficiency on microglial integrity and amyloid pathology in our experimental model, and the compelling evidence that the Y213X mutation causes a marked loss of IL-34 in humans, we next evaluated whether these mechanisms also manifest in human AD cortex. To address this, we performed *postmortem* analyses of temporal cortex tissue from AD patients, examining microglial density, morphology, and tiling in sections from WT individuals and from carriers of the LOF IL-34-Y213X allele. This direct examination of human brain tissue represents an essential step to determine how IL-34 loss shapes microglial responses and amyloid pathology in the human cortex, and to validate the translational relevance of the IL-34-dependent mechanisms uncovered in our study.

Consistent with the graded protein deficiency observed in human carriers, most pronounced in homozygous individuals (≈2 SD lower IL-34 in CSF and ≈3 SD lower in serum; Figure 1), we observed an overall reduction in resident microglia in AD patients homozygous IL-34-Y213X compared to WT individuals, whereas heterozygous carriers showed microglial numbers comparable to WT individuals (p = 0.0008; Figure 6I-J). These data support a requirement for intact IL-34/CSF1R signaling to maintain microglial abundance in humans. In line with IL-34’s role in sustaining gray-matter microglia(*16, 42*), homozygous IL-34 LOF was associated with chronically reduced microglial density. In addition, dispersion index analysis showed that microglia in AD patients homozygous Y213X displayed a more dispersed, non-uniform spatial distribution than WT individuals (p = 0.0093; Figure 6K), indicating altered microglial tissue organization in the setting of IL-34 deficiency.

## Discussion

Our study provides, to our knowledge, the first comprehensive description of human IL-34 deficiency linked to the nonsense variant IL-34-Y213X (p.Tyr213Ter). Across CSF and serum, we observe a dose-dependent reduction in IL-34 that is robust to age, sex, amyloid status and dementia stage, grounding prior genetic associations(*19–21*) in a direct protein phenotype. Population analyses further indicate a geographically structured burden, highest in European and South Asian groups, intermediate in East Asians and lowest or absent in Africans and most admixed American populations.

The marked IL-34 reduction associated with Y213X was consistently detected across orthogonal proteomic platforms (SomaScan in CSF and Olink in serum). AlphaFold modeling(*43, 44*) places the truncation in an unstructured C-terminal region (Supplementary Figure 5), making major disruption of the folded core, and hence of somamer or antibody recognition, unlikely. Nonetheless, we cannot exclude that a truncated IL-34 species is produced but not captured by the detection reagents used; the close phenotypic concordance between IL-34KO mice and homozygous Y213X individuals, particularly the reduction in resident microglial density, argues that any such product would be unlikely to preserve full IL-34 function.

In humans, IL-34 is positioned to shape microglial fitness because it is a high-affinity CSF1R ligand required for maintenance of long-lived, homeostatic microglia in gray-matter territories(*16–18, 42*). Consistent with this biology, low CSF IL-34 co-segregated with coordinated proteomic shifts spanning axon-guidance and synaptic-organization programs and microglial/immune regulators (for example NRXN1, EPHA5, GPNMB and CR1), alongside increases in RNA-binding/splicing proteins, cytokine receptors (including IL21R) and extracellular protease inhibitors (including SERPIN family members), linking IL-34 variation to modules implicated in microglial–neuronal interactions and extracellular remodeling(*10, 16–18, 28, 29*).

Across the AD continuum, CSF IL-34 showed consistent inverse associations with tau-related measures, with stronger effects in carriers and in amyloid-positive individuals, whereas serum IL-34 was only weakly coupled to CSF, supporting compartmentalization and prioritizing CSF IL-34 as a proximal readout of brain IL-34 biology. *In vivo*, IL-34KO mice showed selective depletion of Iba1+ microglia in cortex and hippocampus, consistent with IL-34 as the dominant CSF1R ligand in gray matter(*45, 46*). Importantly, transcriptomic profiling of purified microglia from healthy 9-month-old IL-34KO mice revealed that IL-34 deficiency profoundly reshapes microglial state beyond simply reducing microglial abundance. IL-34KO microglia exhibited a robust pro-inflammatory and disease-associated transcriptional program characterized by induction of canonical DAM and pro-inflammatory microglial mediators together with enrichment of interferon and senescence-associated pathways. Strikingly, these changes were accompanied by impairment of PI3K/AKT/mTOR signaling and oxidative phosphorylation programs, pathways critically involved in maintaining microglial metabolic fitness and homeostatic function (*47–49*). The PI3K/AKT/mTOR axis is a central regulator of microglial energetics and survival, and its disruption has similarly been linked to impaired microglial fitness in TREM2 deficiency (*50*), a well-established AD risk pathway. Concomitant weakening of oxidative phosphorylation further suggests mitochondrial dysfunction and impaired bioenergetic capacity, features commonly associated with dysfunctional and senescent microglia (*48*). Notably, these transcriptional alterations emerged in healthy, unchallenged middle-aged mice lacking overt amyloid pathology or neurodegeneration, indicating that IL-34 deficiency alone is sufficient to promote spontaneous acquisition of inflammatory and neurodegeneration-associated microglial states.

Importantly, IL-34 deficiency also induced dysregulation of several established AD genetic risk factors, including increased expression of *Apoe* and *Clu* together with reduced expression of *Cass4* (*51–54*). The induction of *Apoe*, together with strong enrichment of DAM-associated pathways, is particularly relevant given the central role of APOE-driven microglial reprogramming in AD pathogenesis (*55*). Similarly, altered expressions of *Clu* and *Cass4*, both genetically associated with AD susceptibility, further supports the notion that IL-34 signaling contributes to preservation of a transcriptional network protective against neurodegeneration. Together, these findings position IL-34 upstream of molecular pathways strongly implicated in AD risk and microglial dysfunction.

The cross-species integration provides an additional translational layer linking naturally occurring human IL-34 deficiency to the IL-34KO mouse model. Importantly, the concordance was modest rather than absolute. This is expected given the biological and technical differences between the datasets: human CSF SomaScan measures extracellular and secreted proteins derived from multiple CNS and peripheral sources, whereas the mouse dataset profiles purified microglial RNA. Despite these differences, the significant positive rank-level association suggests that a component of the human IL-34-deficiency proteomic state is recapitulated at the transcriptional level in IL-34-deficient microglia.

Notably, the strongest concordant candidates were not restricted to canonical microglial markers. Instead, they included APLP2, SRXN1, FLT1 and MDGA1, pointing to amyloid-related protein biology, oxidative stress, vascular-inflammatory remodeling and extracellular/synaptic adhesion. This pattern is biologically plausible given the orthogonality of the comparison: CSF SomaScan captures secreted and extracellular proteins from multiple cellular sources, whereas the mouse dataset captures transcriptional responses in purified microglia. The absence of APOE as a direct concordant CSF proteomic signal should not be interpreted as lack of AD relevance; rather, ApoE emerged as a prominent component of the mouse IL-34KO DAM-like transcriptional program, together with Clu induction and Cass4 downregulation.

The strongest signal was observed for proteins increased in human Y213X homozygotes, which tended to align with genes upregulated in IL-34KO microglia, consistent with shared activation of stress, inflammatory, and extracellular remodeling programs. These findings support the use of IL-34KO mice as a mechanistically relevant model of human IL-34 deficiency, while emphasizing that CSF proteomics and microglial transcriptomics provide complementary, not interchangeable, views of IL-34-dependent biology. Thus, rather than implying one-to-one equivalence between human CSF proteins and mouse microglial transcripts, the cross-species analysis supports partial conservation of the molecular consequences of IL-34 loss across systems.

Given the profound transcriptional reprogramming and induction of AD-associated microglial pathways observed in IL-34KO microglia, we next investigated the consequences of IL-34 deficiency in the context of amyloid pathology. IL-34 deficiency in APP/PS1 mice reduced total plaque number, supporting a requirement for homeostatic microglia in early plaque seeding, in line with microglial depletion studies(*36, 56, 57*); residual microglia retained plaque polarization (unlike TREM2 or DAP12 deficiency(*41, 58–60*)) but plaque encapsulation was incomplete and plaques were poorly compacted and dystrophic. Importantly, many of these pathological features are consistent with the transcriptomic phenotype observed in IL-34KO microglia. Senescent and metabolically impaired microglia with defective PI3K/AKT/mTOR signaling and reduced oxidative phosphorylation have been reported to fail in forming an effective protective barrier around plaques, resulting in diffuse and filamentous amyloid structures associated with increased neuritic dystrophy (*48, 61*). Notably, these same features were observed in our APP/PS1 IL-34-deficient mice, supporting a mechanistic link between IL-34-dependent microglial fitness and plaque-associated neuroprotection. Homozygous Y213X individuals similarly exhibited reduced microglial density and disrupted tiling, paralleling the mouse phenotype.

Several limitations warrant consideration, including modest cohort sizes enriched for European ancestry, largely cross-sectional human sampling, restricted postmortem anatomical coverage, and bulk CSF proteomics that do not resolve cell-type specificity. Experimental models necessarily simplify human disease: APP/PS1 captures selected aspects of amyloid pathology, and complete IL-34 loss in mice may not fully mirror lifelong, context-dependent deficiency in human carriers; knock-in strategies approximating Y213X may help bridge these gaps. Finally, additional regulatory variants and pleiotropic associations at the IL-34 locus remain correlative and require functional dissection.

Because Y213X is common in multiple populations, IL-34 deficiency represents a stratifiable biology with non-trivial prevalence. The divergence between CSF and serum supports prioritizing central IL-34 as a pharmacodynamic biomarker for interventions targeting the IL-34/CSF1R axis and provides a framework to test whether restoring IL-34 signaling or modulating downstream microglial state and extracellular programs can alter AD-relevant trajectories in genotype-stratified settings.

## Methods

**Cohort Description.** The ACE Alzheimer Center Barcelona CSF Cohort is part of the Global Neurodegeneration Proteomics Consortium (GNPC), an international collaborative initiative designed to harmonize proteomic biomarker discovery across neurodegenerative diseases(*62*). Participants were recruited through the memory clinic CSF Program, established in 2016 under the IMI2-ADAPTED initiative. The cohort comprises individuals with mild cognitive impairment (MCI) or dementia evaluated at the ACE Memory Clinic, cognitively unimpaired volunteers with subjective cognitive decline (SCD) from the FACEHBI observational study, and early-onset MCI cases enrolled through the BIOFACE study. All participants underwent standardized neurological, neuropsychological, and imaging assessments, with diagnoses established by a multidisciplinary team(*6, 63, 64*).

**Biosample collection.** Cerebrospinal fluid (CSF) was collected under fasting conditions by lumbar puncture according to consensus protocols. Samples were obtained in polypropylene tubes, centrifuged at 2000 × g for 10 minutes at 4 °C, and processed within two hours. On the same day, paired serum, plasma, saliva, and cell pellets were collected, creating a well-characterized biosample repository(*65, 66*).

**Sample characteristics.** The present analysis included n = 1138 participants with CSF IL-34 measurements and n = 461 participants with serum IL-34 measurements, among which n = 459 had paired CSF and serum samples obtained on the same day and under fasting conditions. Dementia status was defined as CDR ≥ 1 (Dementia) versus CDR < 1 (Non-Dementia). Amyloid and tau status were classified according to established biomarker cut-offs within the ACE Alzheimer CSF cohort(*65*). We selected serum aliquots rather than plasma for the measurement of peripheral IL-34, given that serum provides greater stability and reproducibility for immunological mediators and cytokines. Previous studies have shown that cytokines are more reliably preserved in serum, while plasma levels may be strongly influenced by anticoagulants and pre-analytical handling, resulting in higher variability and lower cross-cohort comparability(*67*).

### Array Genotyping and determination of IL-34 Y213X mutation status

DNA samples from the ACE cohort were genotyped using the Axiom 815K Spanish Biobank Array (Thermo Fisher) at the Spanish National Center for Genotyping (CeGEN, Santiago de Compostela, Spain). Genomic quality control (QC) followed standard procedures: samples with low call rate (<97%), excess heterozygosity, sex discrepancies, or relatedness (PI_HAT ≥ 0.1875) were excluded, as were variants with call rate <95% or deviating from Hardy–Weinberg equilibrium (P > 1 × 10⁻⁶). Imputation was performed on the TOPMed server (Michigan, USA), and only variants with MAF > 1%, MAC ≥ 10, and imputation quality R² > 0.3 were retained. Population structure was controlled by principal component analysis (PCA), excluding non-European outliers (>3 SD from the European mean). After QC, 1,259 individuals and 9,016,686 imputed SNPs were available for analysis. The IL-34 Y213X mutation status was determined by extracting the corresponding SNP (rs4985556) from these GWAS data, as previously reported in de Rojas et al.(*19*).

### Somascan analysis of CSF samples

We analyzed 1,370 paired plasma and CSF samples from 1,325 individuals using the aptamer-based high-throughput proteomic platform SomaScan 7k Assay v4.1 (SomaLogic Operating Co., Inc., Boulder, CO). This platform interrogates 7,596 aptamers (“SOMAmers”) corresponding to 6,402 distinct human proteins defined by unique UniProt identifiers. The assay requires only 50 µL of CSF and uses chemically modified DNA aptamers to bind protein targets, with abundance quantified by fluorescence on a DNA array, as originally described by Gold et al.¹¹ Normalization was performed using the adaptive normalization by maximum likelihood (ANML) procedure, as detailed in previous reports and the SomaLogic technical documentation(*68, 69*). Multiple CSF aliquots from the first thaw cycle were used to ensure sample integrity. For downstream analysis, CSF protein intensities were expressed in relative fluorescent units (RFU). Quality control excluded proteins with more than 10% missing values, while remaining missing entries were imputed using column means. RFU values were log-transformed prior to statistical modeling. The complete dataset is available through the GNPC.

**Measurement of IL-34.** CSF IL-34 was quantified using the SomaScan 7.7K platform (Somalogic) with specific SOMAmer reagents targeting interleukin-34 while serum IL-34 was measured using the Olink Explore 3.0 proteomics panel obtained from a previous experiment (Martino-Adami PV et al 2022, Brain). All samples originated from the ACE Alzheimer CSF cohort and consisted of paired CSF and serum samples collected under identical fasting conditions during lumbar puncture.

### Network analysis methods (IL-34 co-regulatory networks)

To investigate co-regulatory protein networks associated with CSF IL-34, we implemented a multi-step pipeline. First, protein abundance data from SomaScan v4.1 (7.7K aptamers) were pre-processed by excluding analytes with more than 10% missing values. For the remaining analytes, missing entries were imputed using the analyte mean. Protein intensity values were then log-transformed to stabilize variance.

To minimize technical confounding, we performed principal component analysis (PCA) on the full proteomic matrix. IL-34 levels and each protein analyte were then residualized against the top principal components (PCs), ensuring that downstream correlations reflected biological rather than technical variation. Next, we computed pairwise Pearson correlations between the residualized IL-34 variable and each residualized protein analyte across the full cohort. For each association, we obtained correlation coefficients and two-sided p-values. Proteins showing an absolute correlation coefficient greater than 0.2 (|r| > 0.2) were retained for downstream network and enrichment analyses. Proteins meeting this threshold were annotated to gene symbols using UniProt identifier mappings. The resulting protein set was then submitted to the STRING database (Search Tool for the Retrieval of Interacting Genes/Proteins) for protein–protein interaction (PPI) analysis. In STRING, we applied a medium-confidence interaction score cutoff of 0.4 and visualized the resulting PPI network. Clustering within the network was performed using the K-means clustering algorithm, and functional enrichment analysis was carried out for Gene Ontology (GO) biological processes and pathways. Enrichment results were corrected for multiple testing (FDR < 0.05).

### Differential expression (DE) analysis methods (IL-34 Y213X vs wild-type)

To examine the proteomic impact of the human IL-34 Y213X mutation in the CSF, we first identified homozygous Y213X carriers and matched controls. Case–control matching was performed using a nearest-neighbor approach. Each homozygous carrier was matched to up to four wild-type controls based on the following variables: age, sex, *APOE* genotype, CSF Aβ42, CSF p-Tau, mini-mental state examination (MMSE) score, and CDR. Matching required at least one *APOE* allele shared between case and controls to ensure genetic comparability. After matching, balance between cases and controls was assessed using Welch’s t-tests (for continuous variables), chi-square tests (for categorical variables), and effect size measures such as standardized mean differences.

For the matched dataset, we performed differential protein expression analysis using ordinary least squares (OLS) regression models for each SomaScan protein analyte. Each model estimated the effect of case–control status (Y213X homozygote vs wild-type) on protein abundance. Analyses were conducted both with and without adjustment for the top two principal components (PC1–PC2) to control for latent technical variation. For each analyte, we obtained regression coefficients, standard errors, t-statistics, and p-values. Proteins with P < 0.05 were considered significantly differentially expressed. Results were visualized with volcano plots, displaying log2 fold changes against –log10 p-values, and top differentially expressed proteins were highlighted. The significant protein sets were further analyzed using the STRING database to identify enriched pathways, GO biological processes, and functional protein clusters.

**Statistical analysis.** Genetic modeling and effect size estimation. The IL-34-Y213X variant (rs4985556) was analyzed both categorically (wild-type, heterozygous, homozygous) and additively using the IL-34_dosage variable (0, 1, or 2 copies of the stop allele). For global genotype associations, we performed linear regression with genotype as a categorical predictor (2 df test). To obtain an objective effect size estimator, we additionally fitted additive dosage models regressing standardized IL-34 levels (CSF and serum separately) on IL-34_dosage. The per-allele regression coefficient (β) was divided by the standard deviation of the outcome to yield a standardized effect size (Cohen’s d). Models were run unadjusted and adjusted for sex, age, principal components (PC1–PC4), and CDR score.

Partial Pearson correlation analyses were performed to evaluate the relationships between IL-34 levels measured in cerebrospinal fluid (CSF; SomaScan assay, target 5400) and serum (Olink platform) with established AD biomarkers (CSF Aβ42, pTau181, and total tau). For each analysis, IL-34 values and biomarker values were residualized with respect to age (Age_LP), sex (Sex_1m_2f), and MMSE (MMSE_PL) using linear regression, and correlations were then computed between the resulting residuals. Subjects were stratified by amyloid status (A+ vs A–, defined by CSF Aβ42 cutoff) and by Y213X mutation carrier status (carriers vs non-carriers). Partial correlation coefficients (r), 95% confidence intervals (derived from Fisher’s z-transformation), and p-values were computed for the full cohort and each subgroup separately. Standardized residuals were visually inspected to confirm model assumptions. Significance thresholds were set at p < 0.05, with more stringent thresholds reported at p < 0.01, p < 0.001, and p < 0.0001.

For other Y213X genotype comparisons, regression models adjusted for sex, age, principal components, and CDR score were applied to estimate adjusted means ± 95% CI, with exact p-values derived from pairwise contrasts. For binary comparisons (sex, dementia, amyloid, tau), group differences were tested by independent t-tests (or non-parametric equivalents when appropriate). Associations with age were assessed using Pearson correlation and linear regression. All analyses were conducted in Python 3.10 using the packages pandas, numpy, statsmodels, and scipy.stats.

### Global allele frequency and distribution analyses of IL-34-Y213X

To characterize the worldwide distribution of the IL-34-Y213X (rs4985556, p.Tyr213Ter) nonsense mutation, we used publicly available allele frequency data from the 1000 Genomes Project Phase 3 (n = 2,504 individuals; 26 populations; 5,008 chromosomes). Data retrieval and processing were performed using the LDpop module of LDlink (v5.0; https://ldlink.nci.nih.gov/), a web-based suite that provides population-specific linkage disequilibrium and allele frequency estimates based on 1000 Genomes reference panels.

For each population, we extracted the observed allele counts and calculated the minor allele frequency (MAF) of rs4985556. Population-specific values were subsequently aggregated into the five continental super-populations defined by the 1000 Genomes Project: AFR (African), AMR (Admixed American), EAS (East Asian), EUR (European), and SAS (South Asian). Global frequencies (ALL) were calculated by pooling across all 26 populations. To estimate the prevalence of IL-34 haploinsufficiency (heterozygous carriers) and IL-34 full deficiency (homozygous carriers), we assumed Hardy–Weinberg equilibrium (HWE) within each super-population. Expected genotype frequencies were derived using the equations: Haploinsufficient (heterozygotes) = 2pq; Full deficiency (homozygotes) = q², where p = frequency of the reference allele and q = frequency of the Y213X stop allele. Carrier rates were expressed as percentages of individuals, while full deficiency rates were additionally reported per 100,000 individuals to facilitate epidemiological interpretation. Two complementary visualizations were generated to summarize the data. First, a geographical allele frequency map was constructed showing the distribution of rs4985556 across all 26 1000 Genomes populations. Each population is displayed at its corresponding geographic location with allele frequencies color-coded on a blue gradient scale. Second, a bar plot of predicted genotype frequencies was produced to illustrate the proportion of haploinsufficient (heterozygous) and fully deficient (homozygous) individuals by continental super-population. Bar colors were pastel shades for clarity, with blue representing haploinsufficiency and red representing full deficiency. Both visualizations are presented together.

### Pleiotropy and regulatory landscape of IL-34 locus

To investigate the broader phenotypic impact of genetic variation at the IL-34 locus, we systematically mined three large-scale resources: (i) the GWAS Catalog (release 2025-05-09), (ii) PheWeb (UK Biobank and FinnGen, release 2025), and (iii) curated datasets of cis-and trans-expression quantitative trait loci (eQTLs) for IL-34. For the GWAS Catalog, we extracted all genome-wide significant associations (P < 5 × 10^−^⁸) reported for SNPs located within or near the IL-34 gene (chr16:66.8–73.2 Mb, GRCh38), as well as associations with distal trans-acting variants reported to influence IL-34 expression. For PheWeb, we retrieved phenome-wide association results for the nonsense variant rs4985556 (Y213X). Data were harmonized by genomic position (GRCh38), rsID, effect allele, trait, and reported p-value. To ensure comparability, traits were grouped into broad physiological systems (brain, eye, systemic, metabolism, morphometry, immune/hematology, etc.) and visualized using heatmaps and system-level bar plots. Finally, cis and trans regulatory loci influencing IL-34 expression were annotated with nearest genes and their functional category, and results were cross-referenced with published references already indexed in the GWAS Catalog.

### Human immunofluorescence

AD samples were provided by Banc de Teixits Neurològics in Biobanc-Hospital Clínic-IDIBAPS, and were processed at the University of Seville. The regional ethical, comité coordinador de ética de la investigación biomédica de andalucía (PID2021-124096BOB-100; 1819-N-22) approved the study. (PID2021-124096BOB-100; 1819-N-22) approved the study. Post-mortem samples from AD patients were received as paraffin-embedded samples. Thus, tissue was first de-paraffinised with xylene for 10 min, followed by rehydration with decreasing ethanol concentrations: 100%, 90%, 70% and 50% until the final distilled water step. Antigen retrieval was applied to the samples to improve antibody linking. Samples were incubated in a 0.1 M sodium citrate solution and heated in a microwave for 5 min. After repeating the previous step, samples were let to recover RT for 20 min. A blocking step consisting of sample incubation in PBS-T 0.05% and 10% BSA was included to improve the signal. Incubation with primary antibody was made overnight in a humidity chamber at 37 °C diluting it in PBS-T 0.05% and 5% BSA until desired concentration. After six washing steps with PBS, we proceeded with secondary antibody (Invitrogen, Thermo Fischer Scientific) incubation diluted 1:300 in PBS-T 0.05% and 5% for 1 h at RT. After that, samples were rinsed three times in PBS; later, samples were immersed in 70% ethanol for 5 min, and autofluorescence was quenched with Autofluorescence Eliminator Reagent (Millipore, Merck) for 5 min; sections were then rinsed in 70% ethanol for three times and 50% glycerol was applied as mounting media, coverslip placed and sealed. Images were obtained from ZEISS LSM 7 DUO confocal microscope (Nikon, Tokyo, Japan). Cell counting of microglial cells was performed using rabbit-derived anti-Ionized calcium-binding adapter molecule 1 (Iba1, 1:500, Fujifilm Wako, Osaka, Japan) and Hoechst 33258 (Invitrogen) at 1:1000 dilution for 5 min as nucleus counterstain. For grey matter determination of microglia, we scanned 1 mm deep from pial surface corresponding to layers I-III using a 20X objective. Next, the Iba1 cells were manually counted in a 1500×1000 µm window. Quantification was made by a blind researcher. To determine the dispersion index, position of each cell within the counting window was saved. Average distance between cells was calculated and divided by the theoretical distance in a totally dispersed counting window. Thus, microglia density was not a factor for dispersion index. Calculations or dispersion index was made in R.

### Animal model

All procedures complied with the European Union Council Guidelines (86/609/EU) and Spanish regulations (BOE 34/11370-421, 2013) for the protection of animals used in research and teaching. Protocols were approved by the Ethical-Scientific Committee of the University of Seville and the Regional Government of Andalusia (Reference code: 06/09/2023/075). Animals were housed at 22 ± 1°C and 60% relative humidity under a 12 h light/dark cycle, with *ad libitum* access to food and water. Both sexes were included to characterize the IL-34KO model in the context of AD.

Four mouse lines were used: APP^swe^, PSEN1^dE9^ (APP/PS1) (The Jackson Laboratory, Bar Harbor, ME, USA, Stock No: 004462), IL-34^em2Smoc^ (Shanghai Model Organisms Center, Shanghai, China. IL-34-KO mice. Stock No: NM-KO-00096) (IL-34KO), and the double mutant IL-34KO-APP/PS1 generated by crossing these strains. Wild-type C57BL/6J mice served as controls. Transgenic animals carried the IL-34 mutation in homozygosity and the APP/PS1 mutation in heterozygosity. The number of animals used is indicated in each graph.

### Intracardiac perfusion and tissue collection

Animals were sacrificed at the predetermined age for tissue collection. Mice were anesthetized with a mixture of ketamine (Ketamidor, 100 mg/ml) and xylazine (Sedin) diluted in 0.9% saline. Intracardiac perfusion was performed using 0.9% saline delivered at a constant flow rate for 4 min using a perfusion pump. Following perfusion, animals were decapitated and brains were rapidly removed. The brains were processed for histology: brains were fixed in 4% PFA for 24 h and then cryoprotected in a sucrose gradient (10%, 20%, 30%). Once tissues sank to the bottom of the tube, they were wrapped in aluminum foil, frozen on dry ice, and stored at –80°C until further processing.

### Cryosectioning

Brain hemispheres prepared for histological analyses were sectioned using a Leica CM1850 UV cryostat at −22°C. Tissue was embedded in OCT compound (Sakura 4583) and mounted onto the cryostat holder. Serial coronal sections (30 μm thickness) were collected for subsequent staining procedures.

### Mice brain immunohistochemistry

Immunohistochemistry was performed to quantify microglia using the Iba1 (1:2000; Wako, 019-19741) marker and to quantify β-amyloid plaques using the Bam10 antibody (1:1000; Anti -β-Amyloid; A3981, Sigma Al). For Iba1 analyses, complete series of sections were collected and quantified in the cortex and the hippocampus. For Bam10 staining, one coronal section per animal containing cortex and hippocampus was analyzed. Sections were washed three times in 1× PBS for 5 min, followed by incubation for 10 min in 99% methanol containing 1% H_2_O_2_ to block endogenous peroxidase activity. Tissue permeabilization was performed with three 5-min washes in 2.5% PBS-T (PBS with 2.5% Triton X-100). To prevent nonspecific binding, sections were incubated for 2 h in 2.5% PBS-T supplemented with 5% BSA. Samples were then incubated overnight at 4°C with the appropriate primary antibody diluted in 2.5% PBS-T with 1% BSA. The following day, sections were equilibrated for 30 min at room temperature and washed three times in 2.5% PBS-T before incubation with the corresponding biotinylated secondary antibody for 2 h. After three washes in 1× PBS, the ABC reagent (Vector PK6100) was applied for 1 h at room temperature.

Visualization was achieved using DAB (Vector SK-4100), and the reaction was stopped with six PBS washes. Sections were mounted on gelatin-coated slides and air-dried overnight. Finally, samples were dehydrated through graded ethanols (70%, 80%, 90%, 100%) and cleared in Histolemon before being coverslipped with DPX for microscopic analysis.

### Mice brain immunofluorescence

Immunofluorescence was performed to visualize antigen-antibody binding using fluorophore-conjugated secondary antibodies, enabling the assessment of marker distribution and potential colocalization. Tissue sections were washed three times in 1× PBS for 10 min, permeabilized in 1% PBS-T for 1 h, and blocked for 1 h in 1% PBS-T containing 5% BSA. Sections were then incubated overnight at 4°C with primary antibodies (Iba1, 1:1000; Wako, 019-19741; Anti-β-Amyloid, 1:1000; A3981, Sigma and Lamp1, 1:250; 1D4B-S DSHB) diluted in 1% PBS-T with 1% BSA. After 24 h, sections were washed six times in 0.1% PBS-T and incubated for 1 h in the dark with the appropriate fluorophore-conjugated secondary antibodies (1:500; Alexa) diluted in 0.1% PBS-T with 1% BSA. When required, fluorescent dye Methoxy-04 (100 µM), ref:863918-78-9 Biotechne Tocris) was added during the final 5 min of this step. Finally, sections were washed six additional times and mounted in a 1:1 PBS–glycerol medium for imaging. Immunofluorescence images were acquired using a Leica Stellaris 8 Falcon confocal microscope (CITIUS I, University of Seville) under identical laser intensity and detector settings for all samples. Subsequent analyses, including quantification and marker colocalization, were performed using Fiji/ImageJ.

### Flow cytometry

We followed the previously described protocol for acute isolation of microglia (n = 6 per genotype) (*70–72*). Mice were anesthetized using pentobarbital (Dolethal) and transcardially perfused with phosphate-buffered saline (PBS). Cortex and hippocampus were dissected out and kept in ice-cold Hank’s Balanced Salt Solution without calcium or magnesium (HBSS, Invitrogen). After mechanical dissociation, the tissue was subjected to enzymatic dissociation using papain (Worthington), at a final concentration of 8 U/mL in combination with DNase I at 80 Kunitz units/ml final concentration (Sigma-Aldrich; D4263) in a piperazine-N,N′-bis (ethanesulfonic acid); 1,4-piperazinediethanesulfonic acid (PIPES)-based buffer with the addition of L-cystein-HCL and ethylenediaminetetraacetic acid (EDTA) under agitation in an incubator at 37 °C for 50 min, and another 15 min of incubation after addition of an extra 25 Kunitz units/ml DNase I. After a 15-minute 200g spin, the pellet was washed and resuspended in Dulbecco’s Modified Eagle Medium (DMEM) with 0.5% bovine serum albumin (BSA) and filtered through a 70-µm single-cell strainer (BD Bioscience). Then, a Percoll gradient was performed (Cytiva, #17089102) at 90% in HBSS (v/v) for microglia and astrocytes enrichment. Cells were stained with primary conjugated monoclonal antibodies CD11b-BV711 (BD bioscience, #563168; 1:200) and CD45-PE (Miltenyi, #130-117-348; 1:200) at 4°C for 30 min. Finally, cells were washed and sorted using MoFlo Astrios (Beckman Coulter) flow cytometers, and data were acquired and analyzed with SUMMIT software (Beckman Coulter). Gating strategy and data analysis were performed in accordance with established guidelines (*73*). Dead cells and debris were excluded based on forward and side scatter profiles. Microglial cells were defined as CD11b⁺CD45⁺ events. Subpopulations were gated using contour density plots set at a 15% probability threshold. Percentages were calculated relative to total single cells.

### Global transcription profiling

Total RNA was extracted from FACS-sorted microglia using TRIzol reagent (Invitrogen), according to the manufacturer’s instructions. RNA integrity and quality were assessed with the Agilent ScreenTape 4200 assay (Agilent). RNA samples were subsequently amplified, and cDNA was generated, hybridized, and stained using the Clariom S Pico Assay for mouse (Applied Biosystems). Arrays were scanned using the Expression Console Software and the GeneChip Scanner 3000 (Affymetrix). Raw data were imported into the R environment and processed using LIMMA/Bioconductor packages in RStudio for quality control, normalization with the Robust Multi-array Average (RMA) method, and differential expression analysis. To investigate biological processes associated with microglia in our mice, we compared WT and IL-34KO microglia using the Gene Set Enrichment Analysis (GSEA) (*74*). Enrichment analysis included Hallmarks gene sets.

### Cross-species integration of human IL-34 deficiency proteomics and mouse microglial transcriptomics

To evaluate whether the proteomic consequences of genetic IL-34 deficiency in humans aligned with the microglial transcriptional response to *IL-34* loss in mice, we performed a cross-species signed-rank integration analysis. Human CSF SomaScan differential expression results from IL-34 Y213X homozygotes versus matched wild-type controls were collapsed to unique gene symbols by retaining the SOMAmer with the strongest nominal association per gene. *IL-34* itself was excluded from cross-species enrichment analyses to avoid circularity.

Human gene symbols were mapped to mouse orthologs, and human proteins were ranked using a signed significance metric defined as sign(coefficient) × −log10(*P* value), where positive values indicate higher abundance in IL-34-deficient individuals. Mouse microglial RNA-seq differential expression results from *IL-34KO* versus wild-type mice were similarly ranked using sign(log2 fold-change) × −log10(*P* value), where positive values indicate higher expression in *IL-34KO* microglia.

Gene set enrichment analysis was performed with fgsea to test whether proteins increased or decreased in human IL-34 deficiency were enriched among genes upregulated or downregulated in *IL-34KO* mouse microglia. In parallel, rank-level concordance across all mapped genes was assessed using Spearman and Pearson correlations between the human and mouse signed-rank statistics. Analyses were performed in R 4.3.3 using data.table and fgsea (*75*).

## Supporting information

Supplementary results

## Data Availability

All data produced in the present study are available upon reasonable request to the authors

## Supplementary Figure Legend

**Supplementary Figure 1.**
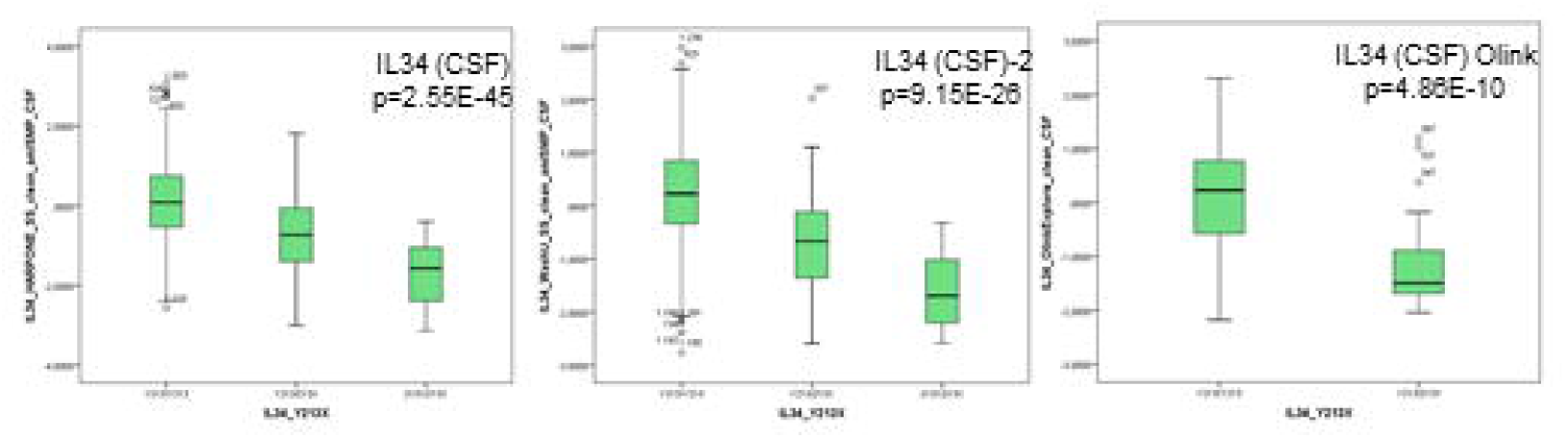
The IL-34-Y213X variant is associated with reduced IL-34 levels in cerebrospinal fluid. Box plots showing CSF IL-34 levels in individuals stratified by IL-34 genotype (Y213/Y213, Y213/X, and X213/X213), measured across three independent experiments using two orthogonal proteomic platforms: two SomaScan assays (left and middle panels) and one Olink immunoassay (right panel). Across all platforms, carriers of the IL-34-Y213X variant show a marked reduction in CSF IL-34 levels. p values indicate statistical significance of genotype effects.

**Supplementary Figure 2.**
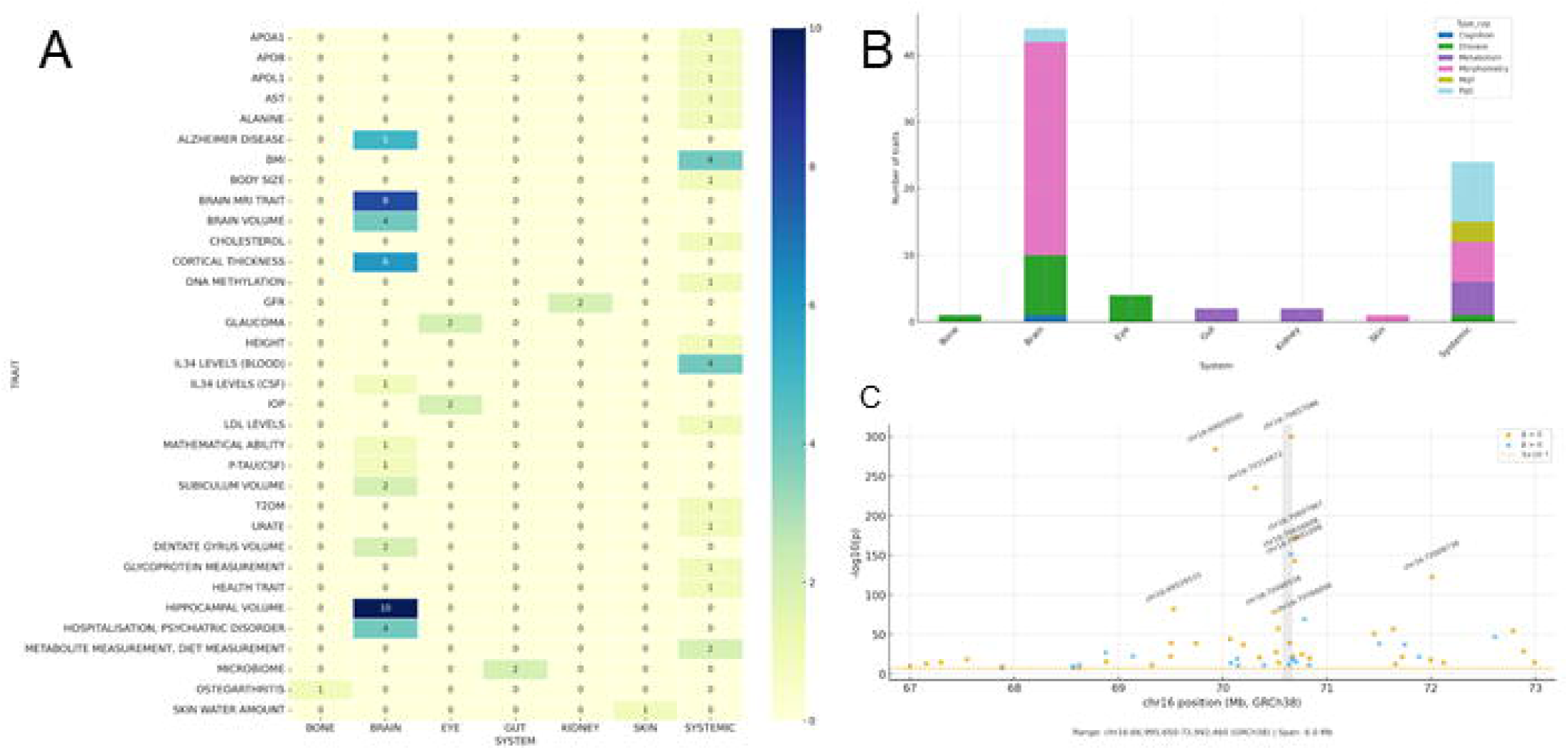
Genome-wide associations of IL-34 locus variants and related phenotypes. (A) Heatmap of genome-wide significant associations for IL-34 locus variants extracted from the GWAS Catalog. Multiple traits reach statistical significance, with strongest signals observed for hippocampal volume, cortical thickness, and brain MRI–derived traits, alongside additional associations with Alzheimer’s disease risk, cognitive performance, mathematical ability, and systemic traits such as APOL1 protein levels. (B) System-level distribution of traits associated with IL-34 locus variants. Pleiotropic effects are evident across multiple biological domains, with a clear predominance in brain- and eye-related phenotypes, but also extending to metabolism, immune/inflammatory traits, and systemic physiology. (C) Regional Manhattan plot of the IL-34 locus (chr16:66.9–73.0 Mb, GRCh38).

**Supplementary Figure 3.**
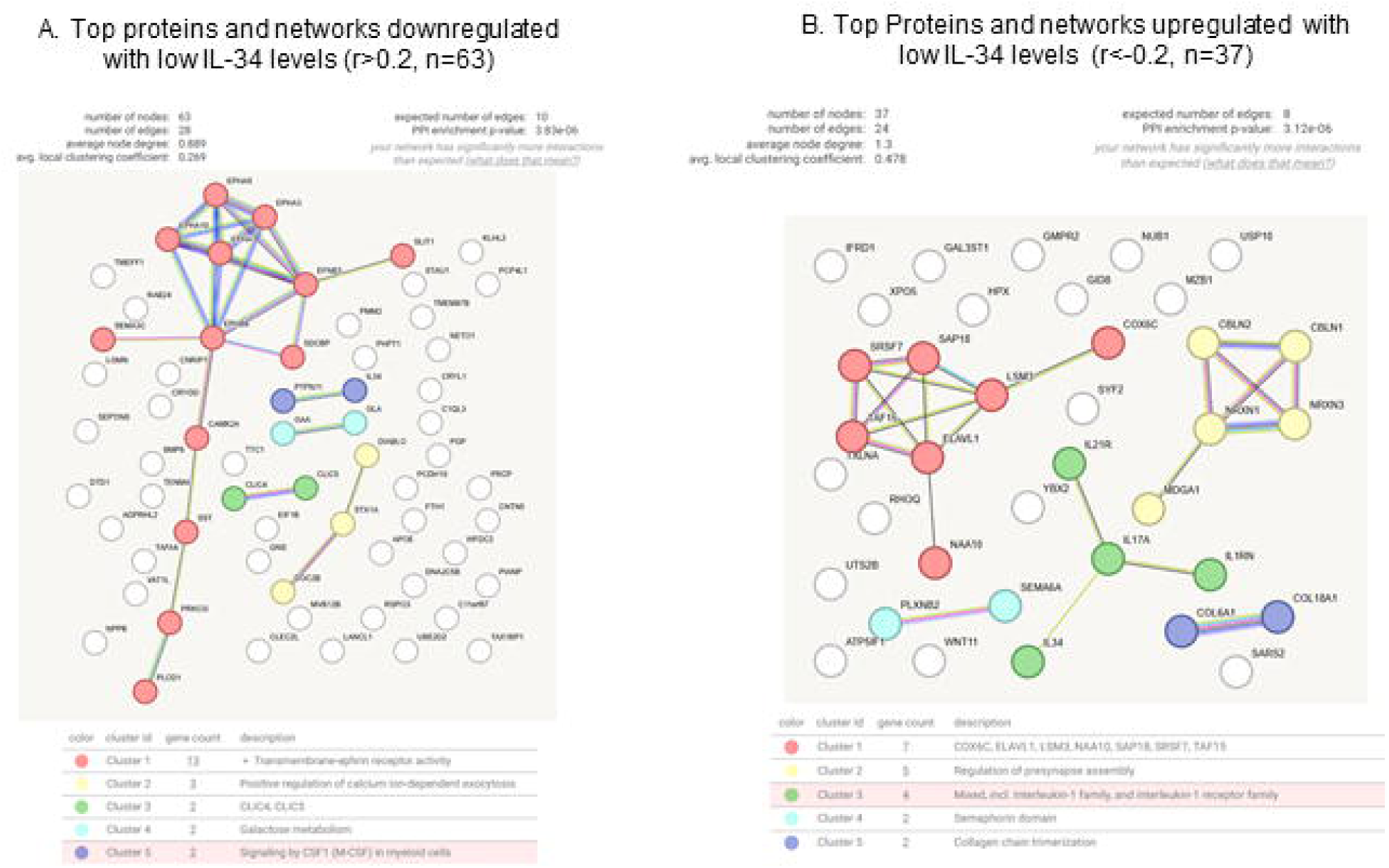
STRING protein–protein interaction network from protein level data and cluster-specific pathway enrichment. STRING PPI network is constructed from proteins with altered levels. Each node represents a protein and edges denote predicted interactions. Node colors indicate cluster membership, with the largest modules corresponding to Cluster 1 (red): Axon guidance, and Cluster 2 (light-orange): Positive regulation of humoral immune response. Smaller modules include galactose metabolism, collagen chain trimerization, and others. Network statistics (inset) confirm significant enrichment over random expectation.

**Supplementary Figure 4.**
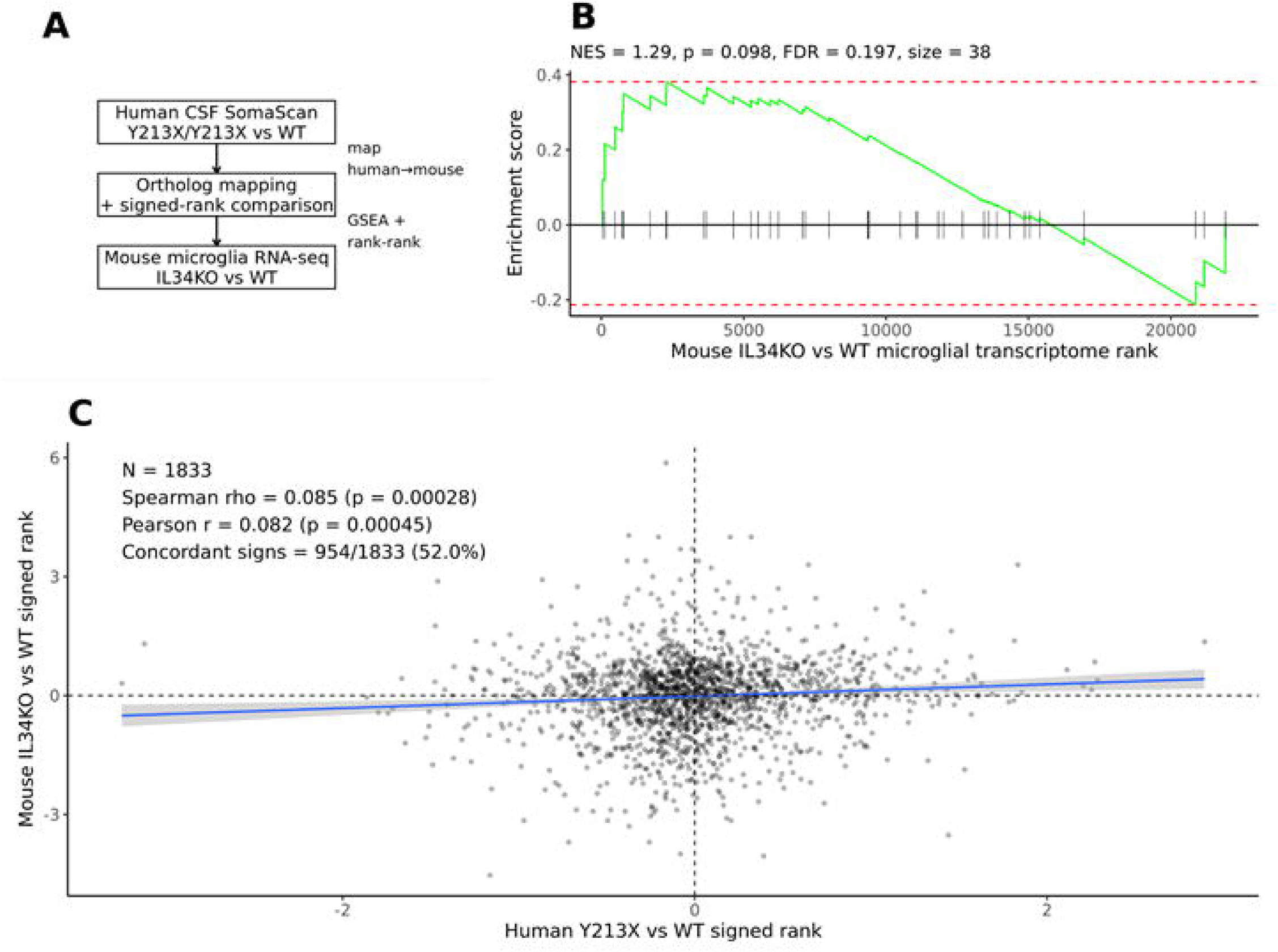
Cross-species comparison of human genetic IL-34 deficiency and IL-34KO mouse microglial transcriptomics. (A) Schematic overview of the cross-species integration strategy comparing the human CSF SomaScan differential signature from IL-34 Y213X homozygotes versus matched wild-type controls with the mouse IL-34KO versus wild-type microglial transcriptome. (B) Gene set enrichment analysis showing the enrichment trend of human proteins increased in IL-34-deficient individuals within the ranked mouse IL-34KO microglial transcriptome. Positive ranks indicate genes increased in IL-34KO microglia. (C) Rank-level comparison across all mapped human–mouse orthologs using signed significance metrics for the human proteomic and mouse transcriptomic contrasts. Positive human ranks indicate increased protein abundance in IL-34 Y213X homozygotes, and positive mouse ranks indicate increased expression in IL-34KO microglia.

**Supplementary Figure 5.**
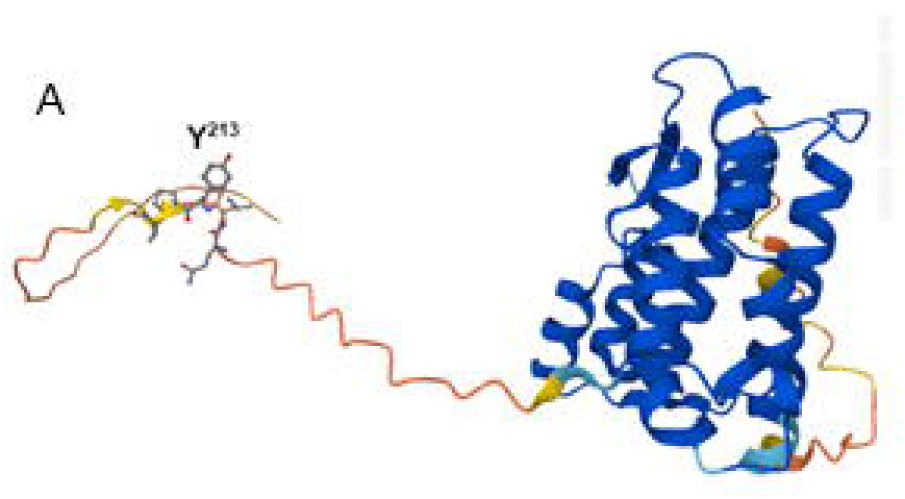
Structural modeling of the IL-34 Y213X truncation. AlphaFold-predicted structure of IL-34 indicating the position of the Y213 residue and the resulting Y213X truncation. The truncation localizes to the unstructured C-terminal region, distal from the folded core of the protein, suggesting that the overall globular structure is preserved and that major disruption of somamer- or antibody-binding epitopes is unlikely.

