## Supplementary table 12 for "Human IL-34 Deficiency Primes Microglia Toward Alzheimer’s Disease-Associated States"

### Supplementary Table 12. Top concordant signals between human IL-34 Y213X CSF proteomics and IL34KO mouse microglial transcriptomics

| **Axis** | **Human protein / gene** | **Human Y213X vs WT** | **Mouse gene** | **Mouse Il34KO vs WT** | **Direction** | **Biological relevance** |
| --- | --- | --- | --- | --- | --- | --- |
| Amyloid / AD-related protein biology | APLP2 | Coef = +7839.8; P = 0.0146 | Aplp2 | log2FC = +0.66; P = 0.0005 | Up / Up | APP-family biology; links IL-34 deficiency to amyloid/neurodegeneration-associated protein remodeling |
| Redox / oxidative stress | SRXN1 | Coef = +33.1; P = 0.0013 | Srxn1 | log2FC = +0.34; P = 0.0442 | Up / Up | Sulfiredoxin-1; supports shared redox-stress response |
| Vascular-inflammatory signaling | FLT1 | Coef = +35.5; P = 0.0498 | Flt1 | log2FC = +0.48; P = 0.0024 | Up / Up | VEGFR1; suggests vascular/inflammatory remodeling under IL-34 deficiency |
| Synaptic / extracellular adhesion | MDGA1 | Coef = +3207.1; P = 0.0349 | Mdga1 | log2FC = +0.32; P = 0.0237 | Up / Up | Synaptic adhesion/extracellular organization; aligns with human IL-34-low CSF network |
| Mitochondrial / stress response | CYCS | Coef = +78.4; P = 0.0054 | Cycs | log2FC = +0.26; P = 0.1422 | Up / Up | Cytochrome c; supports stress/mitochondrial axis, although the mouse signal is not nominally significant |
| Glycosylation / extracellular plasticity | GALNT14 | Coef = +92.4; P = 0.0052 | Galnt14 | log2FC = +0.21; P = 0.4194 | Up / Up | Protein glycosylation/extracellular remodeling; human signal strong, mouse direction concordant |
| Neurodevelopment / synaptic signaling | NRG2 | Coef = +15.9; P = 0.0270 | Nrg2 | log2FC = +0.45; P = 0.1044 | Up / Up | Neuregulin signaling; suggests shared neuroglial/synaptic remodeling |
| Glutamate/peptidase-related biology | NAALAD2 | Coef = +4.0; P = 0.0153 | Naalad2 | log2FC = +0.22; P = 0.0414 | Up / Up | Enzymatic/extracellular signaling; concordant and nominal in both datasets |
| TNF receptor / injury signaling | TNFRSF21 | Coef = -4777.2; P = 0.0325 | Tnfrsf21 | log2FC = -0.58; P = 0.0179 | Down / Down | TNF receptor family member; suggests selected immune/injury signaling is reduced in both systems |
| Immune cytoskeleton / myeloid signaling | HCLS1 | Coef = -73.8; P = 0.0426 | Hcls1 | log2FC = -0.48; P = 0.0262 | Down / Down | Hematopoietic cell-specific Lyn substrate; immune/cytoskeletal signaling |
| Notch/glycosylation | POFUT1 | Coef = -99.5; P = 0.0227 | Pofut1 | log2FC = -0.26; P = 0.0633 | Down / Down | Protein O-fucosylation/Notch-related biology; trend in mouse |
| Endosomal / ubiquitin signaling | STAMBP | Coef = -6.45; P = 0.0297 | Stambp | log2FC = -0.78; P = 0.0861 | Down / Down | Endosomal/deubiquitination biology; concordant direction |

*Note: Values are provided for the concordant signals highlighted in the cross-species comparison. Positive human coefficients indicate higher CSF protein abundance in IL-34 Y213X homozygotes; positive mouse log2FC values indicate higher expression in Il34 knockout microglia.*
