## Supplementary results for "Human IL-34 Deficiency Primes Microglia Toward Alzheimer’s Disease-Associated States"

1. **Cis-regulatory and trans regulatory variants modulates peripheral IL-34 plasma level expression**

To identify additional variation modulating IL-34 cytokine expression, we first examined cis-pQTLs for IL-34 on chromosome 16 reported in GWAs catalog (Supplementary Table S7). Multiple highly significant local signals were identified in IL-34 plasma levels. The strongest association mapped near exon 5 of IL-34 (chr16:70657086, rs8046424; p = 1.0 × 10^-^³⁰⁰), which was linked to a 0.42 unit decrease in circulating IL-34 levels. Additional independent peaks included chr16:69929500 (rs1875245, WWP2; p = 9.0 × 10^-^²⁸⁵; β = –0.71) and chr16:70314872 (DDX19B; p = 8.0 × 10^-^²³⁶; β = –0.89), both associated with marked protein reduction (Supplementary results Figure 1C). Several other named SNPs also exerted strong effects. For example, rs3785423-T (chr16:70.698 Mb, VAC14) decreased IL-34 by 0.46 units (p = 4.0 × 10^-^¹⁷³), whereas rs118062333-C (chr16:70.656 Mb, IL-34) was associated with a 1.52 unit increase in IL-34 (p = 1.0 × 10^-^¹⁵¹). Together, these results indicate that IL-34 expression is under the control of multiple independent cis-regulatory variants spanning 6 Mb.

Beyond local regulation, several *trans* loci reached genome-wide significance for IL-34 protein levels (Supplementary Table S7). The most significant trans-acting signal was detected at the CFH locus on chromosome 1 (rs34813609; p = 1.0 × 10^-^¹⁷⁴), associated with a 0.37 unit decrease in circulating IL-34. A second major peak mapped to the complement component 2 (C2) region on chromosome 6 (chr6:31943585-T, rs557109896; p = 2.0 × 10^-^⁴³), while a third strong association was identified at the BCHE/LINC01322 locus on chromosome 3 (rs71674639; p = 5.0 × 10^-^²¹). Additional signals included rs35267984 (MED16, chr19; p = 2.0 × 10^-^¹⁹), rs74480769-G (C7, chr5; p = 2.0 × 10^-^¹⁵), and rs12318001-C (CHST11, chr12; p = 8.0 × 10^-^¹⁵). Strikingly, multiple trans associations mapped to complement-related genes (CFH, C2, C7), reinforcing the involvement of complement activation pathways in IL-34 regulation. This is consistent with the protein co-expression networks observed in CSF and with differential protein expression patterns detected when comparing Y213X homozygotes against wild-type individuals. Together, these results highlight a strong link between IL-34 biology and systemic immune processes, particularly complement activation and extracellular matrix regulation.

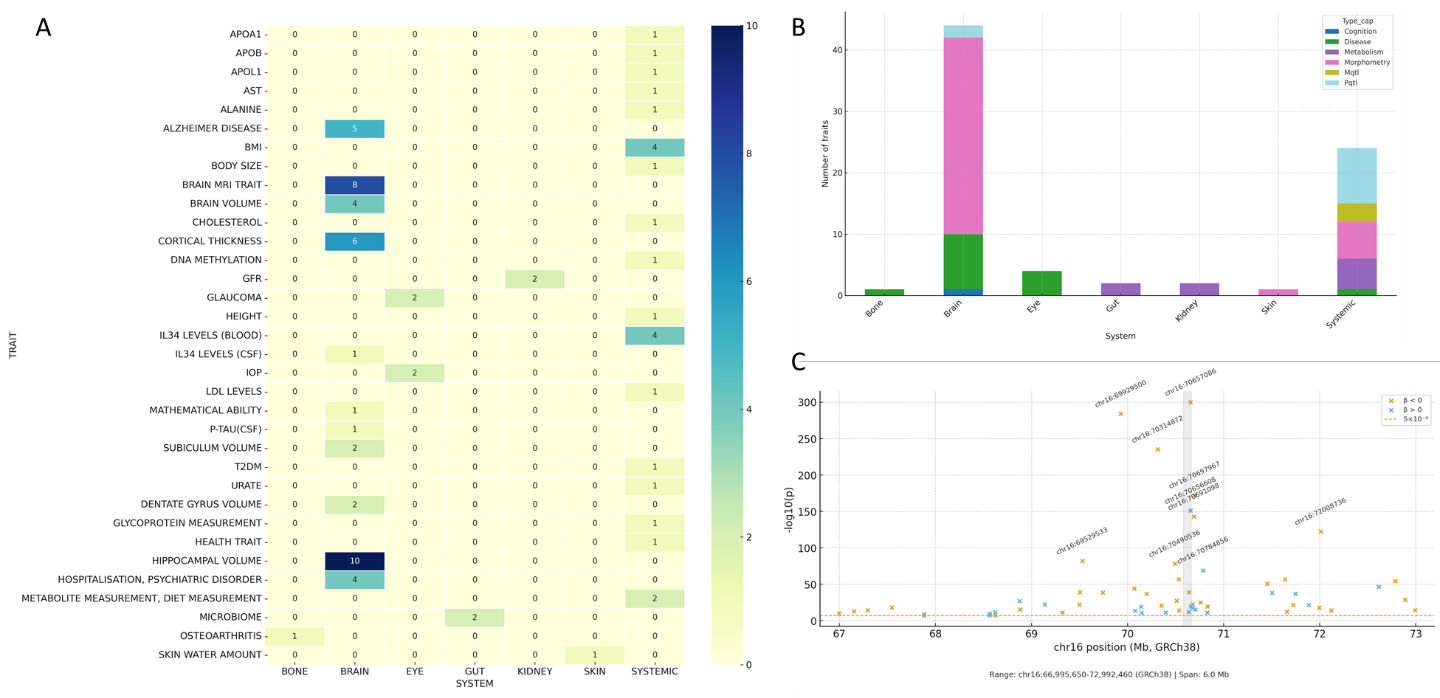

**Supplementary Figure 2: Genome-wide associations of IL-34 locus variants and related phenotypes.** (A) Heatmap of genome-wide significant associations for IL-34 locus variants extracted from the GWAS Catalog. Multiple traits reach statistical significance, with strongest signals observed for hippocampal volume, cortical thickness, and brain MRI–derived traits, alongside additional associations with Alzheimer’s disease risk, cognitive performance, mathematical ability, and systemic traits such as APOL1 protein levels. (B) System-level distribution of traits associated with IL-34 locus variants. Pleiotropic effects are evident across multiple biological domains, with a clear predominance in brain- and eye-related phenotypes, but also extending to metabolism, immune/inflammatory traits, and systemic physiology. (C) Regional Manhattan plot of the IL-34 locus (chr16:66.9–73.0 Mb, GRCh38).

2. **Additional phenotypic associations within IL-34 gene.**

Mining of the GWAS Catalog revealed that *IL-34* locus variation contributes to a wide spectrum of human phenotypes beyond cytokine levels (Supplmentary Figure 1A–B). Within the CNS, variants were strongly associated with neuroimaging-derived traits, including brain volume, cortical thickness, hippocampal subfields (subiculum and dentate gyrus), and other MRI-derived morphometric measures (p values ranging from 10^-^⁵⁶ to 10^-^⁹) and with psychiatric-related outcomes such as hospitalisation for psychiatric disorders (p = 4.0 × 10^-^⁶), pointing to a role for IL-34-linked pathways in neurodegenerative and neuropsychiatric conditions (Supplementary table 8).

Beyond the brain, ocular traits such as glaucoma (p = 2.0 × 10^-^¹¹) and intraocular pressure (p = 7.0 × 10^-^⁹) reached genome-wide significance, consistent with shared biology between retinal microglia and central microglial populations. Systemically, IL-34 locus variation was also associated with metabolic and cardiovascular phenotypes, including BMI (p = 3.0 × 10^-^¹²), GFR (p = 2.0 × 10^-^¹⁵), urate, glycoprotein and cholesterol levels, as well as circulating apolipoproteins (APOA1, APOB, APOL1; P values 10^-^¹³-10^-^¹⁰). Additional associations encompassed osteoarthritis (p = 4.0 × 10^-^¹⁶), type 2 diabetes (p = 4.0 × 10^-^⁸), and systemic metabolic traits such as alanine and DNA methylation, underscoring the pleiotropic effects of this locus. Together, these findings indicate that IL-34-linked variants are not only central to Alzheimer’s disease, microglial and brain structural phenotypes but also extend their impact to systemic metabolic regulation, immune-related processes, and ocular health. This pleiotropy suggests that IL-34 biology bridges neuroimmune regulation with systemic homeostasis, offering a potential mechanistic link between microglial pathways, complement activity, and whole-body metabolic traits.
